# Association of Genetic Ancestry with Molecular Tumor Profiles in Colorectal Cancer

**DOI:** 10.1101/2023.07.12.23292571

**Authors:** Brooke Rhead, David M. Hein, Yannick Pouliot, Justin Guinney, Francisco M. De La Vega, Nina N. Sanford

## Abstract

**Background:** Prior research on molecular correlates of disparities in incidence and outcomes of colorectal cancer (CRC) have typically used self-reported or observed categories of race and ethnicity, which can be missing or inaccurate. Furthermore, race and ethnicity do not always capture genetic similarity well, particularly in admixed populations. To overcome these limitations, we examined associations of CRC tumor molecular profiles using genetic ancestry.

**Methods:** Sequencing was performed with the Tempus xT NGS 648-gene panel and whole exome capture RNA-Seq for 8,454 CRC patients. Genetic ancestry proportions were estimated for five continental groups, Africa (AFR), Americas (AMR), East Asia (EAS), Europe (EUR), and South Asia (SAS), using ancestry informative markers. We assessed association of genetic ancestry proportions and genetic ancestry-imputed race and ethnicity categories with somatic mutations in relevant CRC genes and in expression profiles, including consensus molecular subtypes (CMS).

**Results:** Increased AFR ancestry was associated with higher odds of somatic mutations in *APC*, *KRAS* and *PIK3CA* and lower odds of *BRAF* mutations. Additionally, increased EAS ancestry was associated with lower odds of mutations in *KRAS*, EUR with higher odds in *BRAF*, and the Hispanic/Latino category with lower odds in *BRAF*. Greater AFR ancestry and the non-Hispanic Black category were associated with higher rates of CMS3, while patients in the Hispanic/Latino category had more indeterminate CMS.

**Conclusions:** Use of genetic ancestry enables identification of molecular differences in CRC tumor mutation frequencies and gene expression that may underlie observed differences by race and ethnicity, and suggests that subtype classifications such as CMS may benefit from greater patient diversity.

## Background

Overall incidence and mortality of colorectal cancer (CRC) has declined over the last several decades due to a combination of risk reduction, early detection, and advancements in therapy.^1^ However, there has been a growing burden of CRC among young adults and persistent disparities in outcomes by race and ethnicity across all ages.^2^ As such, improved CRC outcomes are not equally realized across demographics in the United States.

The rising incidence of CRC among adults aged <50 years, termed early onset CRC (EOCRC), has garnered significant attention by patients, media, and clinicians. Patients with EOCRC typically have delayed presentation, leading to more advanced disease at time of diagnosis.^3^ To date, studies have not demonstrated consistent, clinically relevant molecular differences in early versus average onset CRC (AOCRC).^4–6^ As such, the cause for increasing incidence of EOCRC is largely attributed to potential environmental and behavioral components, with specific factors yet to be elucidated.^3^

Racial and ethnic differences in CRC outcomes are also multifactorial in etiology. Longstanding disparities in access to care have disproportionately affected Black populations who have the worst CRC outcomes, regardless of clinical factors such as age or stage at diagnosis.^7–9^ Prior studies have also demonstrated molecular differences in CRC by race and ethnicity with predictive and prognostic implications, including increased prevalence of *KRAS* mutations among Black patients.^8,10–12^ However, most of these studies use self-reported or observed race and ethnicity categories that are often either missing or imprecise, particularly in real-world data and in highly admixed groups such as Black and Hispanic/Latino patients.^13^ In contrast, genetic ancestry, assessed via a patient’s sequencing or genotyping data, can potentially better capture genotypic profiles associated with risk and provide more accurate information for population stratification.^14^

Given disparities in incidence and outcome of CRC by race, ethnicity, and age, along with the limitations of traditionally used race and ethnicity categories based on the US government’s Office of Management and Budget standard,^15^ we examined whether genetic ancestry proportions were associated with patterns of molecular alterations in CRC using a large, real-world cohort. To address the missingness of race and ethnicity data common in real-world datasets, we imputed these categories from genetic ancestry. We then evaluated associations with the imputed categories, both to compare to our genetic ancestry proportion findings and to prior research using self-reported categories. Furthermore, we assessed whether race and ethnicity associations were different in AOCRC versus EOCRC, or by primary tumor site.

## Methods

### Patient cohort

Genomic and clinical metadata of 8,454 patients diagnosed with CRC were obtained from the Tempus database, which includes de-identified genomic and clinical data from cancer patients that underwent tumor profiling as part of their healthcare. Selection criteria included tumor profiling with the Tempus xT assay(v2-v4) from 2018 to 2022. Briefly, the assay is a targeted panel that detects single nucleotide variants, insertions and/or deletions, and copy number variants in 598-648 genes, as well as chromosomal rearrangements in 22 genes with high sensitivity and specificity. A subset of those patients had additional whole exome RNA sequencing.^16^ Available demographic information included: patient age at date of specimen collection, age at diagnosis, gender, stated (i.e., either self-reported or observed) race and ethnicity, and smoking status. Primary tumor site in addition to clinical details such as tumor grade, microsatellite instability (MSI) status, and sequenced tissue site were included. Analysis was performed using de-identified data under the exemption Pro00042950 granted by Advarra, Inc. Institutional Review Board (IRB).

### CRC-relevant genes and mutation types

Genes relevant to CRC were identified from the following sources: 187 genes belonging to 10 oncogenic signaling pathways reported by Sanchez-Vega et al.,^26^ 72 genes predicted by the Integrative OncoGenomics pipeline to be CRC drivers (IntOGen, release date 2020.02.01),^27^ 15 genes associated with hereditary colorectal cancer syndromes (Lynch, Li Fraumeni, and polyposis syndromes) for which germline variants are reportable in the Tempus xT assay, and 22 genes that were investigated in a previous CRC study that utilized Tempus data.^10,16^ Of these genes, 137 are included in the Tempus xT assay gene panels (v2-v4).

Different mutation types were evaluated: (1) protein-altering somatic mutations, defined as single or multiple nucleotide mutations, short insertions or deletions (<=50 bp) and other changes that impact protein structure or splice sites (Sequence Ontology, SO:0001818), (2) somatic copy number alterations (SCNAs), defined as structural insertions or deletions greater than 500 bp in size, and (3) actionable mutations, defined in our study as protein-altering mutations with an OncoKB Therapeutic Level of Evidence V2 designation of therapeutic level 1 or 2, or resistance level R1, irrespective of the type of solid cancer.^28^ For protein-altering mutations, only patients with matched normal tissues were included in analyses due to the potential for germline variants to be misclassified as somatic when normal tissue is unavailable. Patients without matched normal tissues were included in analyses of SCNAs and actionable mutations. We required a prevalence of at least 1% (and minimum 10 patients) for a specific mutation type to include a gene for evaluation.

### Determination of genetic ancestry

Genetic ancestry proportions were estimated using a supervised global genetic ancestry estimation algorithm.^17^ Proportions for five continental ancestry groups – Africa (AFR), the Americas (AMR), East Asia (EAS), Europe (EUR), and South Asia (SAS) – were calculated using 654 ancestry informative markers (AIMs) that overlap targeted regions of the Tempus xT NGS assay.^16,18^ Reference allele frequency data for the AIMs was obtained from the 1,000 Genomes Project,^19^ The Human Genome Diversity Project,^20^ and the Simons Genome Diversity Project databases.^21^ The accuracy of our methods was evaluated using published ancestry proportions determined using the gold standard method, RFMix,^22^ on whole-genome sequencing data from the Pan-Cancer Analysis of Whole Genomes Project.^23^ Normal specimens were used to determine genotypes at the AIMs when available; otherwise tumor data were used.

### Association between genetic ancestry and somatic mutations

Likelihood ratio tests (LRTs) were used to identify genes in which the presence of somatic mutation was associated with genetic ancestry proportions or race and ethnicity imputed categories. For each gene, a multivariable logistic regression model that included somatic mutation (presence/absence) or copy number alteration as dependent variables, and ancestry proportions, assay version, gender, and age at sample collection as independent variables (full model) was compared to a nested model that excluded ancestry proportions. LRT p-values were corrected for the number of genes tested using the Benjamini-Hochberg method. For genes with significant LRT p-values, specific genetic ancestry proportion associations (AMR, AFR, EAS, EUR or SAS) were identified in the full model (any uncorrected coefficient p<0.05 considered significant).

In order to include all five genetic ancestry proportions in the same model, so that each ancestry association was adjusted for the remaining four ancestries while also properly accounting for data compositionality, proportions were first transformed into an isometric log ratio (ILR) representation (“pivot coordinates”) using the pivotCoord function in the robCompositions R package.^29^ Analyses were then repeated using imputed race and ethnicity categories in place of ancestry proportions, with “complex” excluded from further analyses, and the NH White group used as the reference category. Tests were stratified by microsatellite instability (MSI) status as determined by the Tempus xT algorithm. Odds ratios (ORs) and 95% confidence intervals were estimated from the full models. Complete case analysis was utilized in all regression models.

In addition to the main analyses, we looked for imputed race and ethnicity category associations that differed by age of diagnosis (EOCRC vs. AOCRC), or by cancer primary site sidedness (colon vs. rectum and left colon/rectum vs. right colon) among microsatellite stable (MSS) patients. To identify such associations, we first conducted LRTs with logistic models similar to those in the main analyses, but with added indicator variables for age of diagnosis (or cancer primary site) in both the full and nested models, and an interaction term for age of diagnosis (or cancer primary site) and imputed race and ethnicity category in the full model. LRT p-values were corrected for multiple hypotheses using the Benjamini-Hochberg method. For any genes where evidence of interaction was identified by the LRT, interaction terms in the full model with p<0.05 identified specific race and ethnicity categories with interaction effects. The full models were used to estimate ORs and 95% confidence intervals.

Among patients with MSI-high status, the Kruskal-Wallis test was used to assess whether there were differences in tumor mutation burden (TMB, number of mutations/megabase) by age at diagnosis (AOCRC vs. EOCRC), by imputed race and ethnicity alone, and by imputed race and ethnicity stratified by age at diagnosis.

### Imputation of race and ethnicity categories

To overcome missingness of stated race and ethnicity in our real-world data (cf. Table 1), imputation of mutually exclusive race and ethnicity categories from genetic ancestry proportions were estimated using a set of heuristics (Supplementary Table S1) derived from admixture proportions reported in the literature for Black and Hispanic/Latino groups in the United States.^24^ Four categories were defined, non-Hispanic (NH) Asian, NH Black, Hispanic/Latino, and NH White, with patients remaining unclassified under our heuristics termed “complex”. Cutoffs for category membership were additionally empirically adjusted to better reflect genetic ancestry proportions observed in patients with available self-reported or observed race and ethnicity data. The assessment of the sensitivity and specificity of our imputation method, as shown in Supplementary Table S1, demonstrates high accuracy in our dataset and enabled us to use this data for comparisons across categories.

**Table 1:** Patient characteristics by imputed race and ethnicity category.

| **Characteristic** | **NH White,**<br>**N = 5,763<sup>1</sup>** | **NH Black,**<br>**N = 1,061<sup>1</sup>** | **NH Asian,**<br>**N = 420<sup>1</sup>** | **Complex,**<br>**N = 258<sup>1</sup>** | **Hispanic/**<br>**Latino,**<br>**N = 952<sup>1</sup>** | **p-value<sup>2</sup>** |
| --- | --- | --- | --- | --- | --- | --- |
| **Stated race** |  |  |  |  |  | <0.001 |
| White | 3,268<br>(97%) | 23 (3.4%) | 9 (4.1%) | 95 (78%) | 223 (57%) |  |
| American Indian or<br>Alaska Native | 2 (<0.1%) | 0 (0%) | 3 (1.4%) | 3 (2.5%) | 29 (7.4%) |  |
| Asian | 2 (<0.1%) | 0 (0%) | 181 (83%) | 10 (8.2%) | 1 (0.3%) |  |
| Black or African<br>American | 9 (0.3%) | 634 (94%) | 0 (0%) | 0 (0%) | 6 (1.5%) |  |
| Native Hawaiian or<br>Other Pacific Islander | 1 (<0.1%) | 0 (0%) | 4 (1.8%) | 2 (1.6%) | 1 (0.3%) |  |
| Other Race | 75 (2.2%) | 20 (3.0%) | 19 (8.8%) | 12 (9.8%) | 129 (33%) |  |
| Race not stated | 6 (0.2%) | 0 (0%) | 1 (0.5%) | 0 (0%) | 1 (0.3%) |  |
| Unknown | 2,400 | 384 | 203 | 136 | 562 |  |
| **Stated ethnicity** |  |  |  |  |  | <0.001 |
| Not Hispanic or Latino | 1,717<br>(98%) | 277 (95%) | 120 (99%) | 64 (75%) | 67 (14%) |  |
| Hispanic or Latino | 37 (2.1%) | 14 (4.8%) | 1 (0.8%) | 21 (25%) | 414 (86%) |  |
| Unknown | 4,009 | 770 | 299 | 173 | 471 |  |
| **Age at specimen**<br>**collection** | 62 (52, 70) | 60 (50, 68) | 60 (50, 68) | 59 (50, 69) | 56 (47, 66) | <0.001 |
| Unknown | 10 | 1 | 0 | 0 | 3 |  |
| **Age at onset** | 60 (51, 69) | 58 (49, 67) | 58 (49, 66) | 57 (48, 67) | 55 (45, 64) | <0.001 |
| Unknown | 1,003 | 185 | 64 | 47 | 142 |  |
| **Onset age group** |  |  |  |  |  | <0.001 |
| AOCRC | 3,664<br>(77%) | 635 (72%) | 255 (72%) | 153 (73%) | 514 (63%) |  |
| EOCRC | 1,096<br>(23%) | 241 (28%) | 101 (28%) | 58 (27%) | 296 (37%) |  |
| Unknown | 1,003 | 185 | 64 | 47 | 142 |  |
| **Gender** |  |  |  |  |  | 0.4 |
| Female | 2,455<br>(43%) | 487 (46%) | 179 (43%) | 115 (45%) | 420 (44%) |  |
| Male | 3,287<br>(57%) | 572 (54%) | 239 (57%) | 143 (55%) | 528 (56%) |  |
| Unknown | 21 | 2 | 2 | 0 | 4 |  |
| **xT assay version** |  |  |  |  |  | 0.3 |
| xT.v2 | 857 (15%) | 149 (14%) | 68 (16%) | 36 (14%) | 120 (13%) |  |
| xT.v3 | 1,076<br>(19%) | 204 (19%) | 64 (15%) | 44 (17%) | 194 (20%) |  |
| xT.v4 | 3,830<br>(66%) | 708 (67%) | 288 (69%) | 178 (69%) | 638 (67%) |  |
| **Smoking status** |  |  |  |  |  | <0.001 |
| Never smoker | 2,007<br>(51%) | 392 (55%) | 184 (67%) | 86 (55%) | 375 (60%) |  |
| Ever smoker | 1,893<br>(49%) | 323 (45%) | 89 (33%) | 70 (45%) | 255 (40%) |  |
| Unknown | 1,863 | 346 | 147 | 102 | 322 |  |
| **Cancer stage** |  |  |  |  |  | 0.019 |
| Stage 1 | 23 (0.6%) | 7 (1.0%) | 1 (0.4%) | 2 (1.3%) | 5 (0.9%) |  |
| Stage 2 | 163 (4.6%) | 32 (4.6%) | 7 (2.8%) | 11 (7.1%) | 24 (4.2%) |  |
| Stage 3 | 444 (12%) | 89 (13%) | 40 (16%) | 21 (14%) | 107 (19%) |  |
| Stage 4 | 2,925<br>(82%) | 568 (82%) | 202 (81%) | 120 (78%) | 439 (76%) |  |
| Unknown | 2,208 | 365 | 170 | 104 | 377 |  |
| **Tumor grade** |  |  |  |  |  | 0.002 |
| Low | 361 (9.5%) | 56 (7.9%) | 30 (10%) | 7 (4.5%) | 78 (12%) |  |
| Medium | 2,549<br>(67%) | 511 (72%) | 199 (69%) | 110 (70%) | 474 (71%) |  |
| High | 899 (24%) | 144 (20%) | 61 (21%) | 40 (25%) | 117 (17%) |  |
| Unknown | 1,954 | 350 | 130 | 101 | 283 |  |
| **MSI status** |  |  |  |  |  | 0.062 |
| Low/Stable | 5,432<br>(94%) | 1,017 (96%) | 405 (96%) | 247 (96%) | 896 (94%) |  |
| High | 322 (5.6%) | 42 (4.0%) | 15 (3.6%) | 10 (3.9%) | 56 (5.9%) |  |
| Unknown | 9 | 2 | 0 | 1 | 0 |  |
| **TMB count** | 3 (2, 5) | 4 (2, 6) | 3 (2, 5) | 3 (2, 5) | 3 (2, 5) | <0.001 |
| Unknown | 4 | 2 | 1 | 0 | 0 |  |
| **Tumor/normal tissue status** |  |  |  |  |  | 0.022 |
| Tumor and normal | 3,486<br>(61%) | 659 (62%) | 246 (59%) | 152 (59%) | 626 (66%) |  |
| Tumor only | 2,270<br>(39%) | 401 (38%) | 174 (41%) | 105 (41%) | 326 (34%) |  |
| Unknown | 7 | 1 | 0 | 1 | 0 |  |
| **Cancer primary site** |  |  |  |  |  | <0.001 |
| Left colon | 524 (9.1%) | 88 (8.3%) | 52 (12%) | 34 (13%) | 98 (10%) |  |
| Not specified | 3,823<br>(66%) | 712 (67%) | 253 (60%) | 153 (59%) | 609 (64%) |  |
| Rectum | 850 (15%) | 128 (12%) | 72 (17%) | 46 (18%) | 161 (17%) |  |
| Right colon | 566 (9.8%) | 133 (13%) | 43 (10%) | 25 (9.7%) | 84 (8.8%) |  |
| **Sequenced tissue site** |  |  |  |  |  | <0.001 |
| CRC (primary) | 2,841<br>(49%) | 556 (53%) | 211 (50%) | 137 (53%) | 554 (58%) |  |
| Liver | 1,270<br>(22%) | 242 (23%) | 75 (18%) | 47 (18%) | 185 (19%) |  |
| Lung | 347 (6.0%) | 41 (3.9%) | 39 (9.3%) | 11 (4.3%) | 43 (4.5%) |  |
| Other | 1,285<br>(22%) | 220 (21%) | 95 (23%) | 62 (24%) | 168 (18%) |  |
| Unknown | 20 | 2 | 0 | 1 | 2 |  |

<sup>1</sup> n (%); Median (IQR)
<sup>2</sup> Fisher's Exact Test for Count Data with simulated p-value (based on 2000 replicates) for categorical variables with any expected cell count <5; Pearson's Chi-squared test for categorical variables with all expected cell counts ≥5; Kruskal-Wallis rank sum test for continuous variables

### Differences in cohort characteristics by imputed race and ethnicity category

Differences in cohort characteristics among imputed race and ethnicity categories were assessed using the R package *gtsummary*.^25^ Fisher’s exact test for count data with simulated p-value (based on 2000 replicates) was used for categorical variables with any expected cell count <5, Pearson’s Chi-squared test was used for categorical variables with all expected cell counts ≥5, and the Kruskal-Wallis rank sum test was used for continuous variables.

### Gene expression data exploration and preparation

Tempus xT RNA-Seq raw sequencing data were processed with Kallisto to quantify transcript abundances as previously described.^30^ Raw transcript counts were filtered to a minimum of 10 counts in 5% of samples and a variance stabilized transform (VST, DESeq2) was applied.^31^ Batch effects due to assay version were assessed with Principal Component Analysis (PCA) and removed with LIMMA via linear modeling (removeBatchEffect).^32^ PCA plots labeled by grade, MSI status, tissue site, and clinical stage were then generated and inspected for the presence of clustering and used to inform subsetting of patients for separate downstream testing. Further variable selection for multivariable analyses was then performed on each subset. First, variables with more than 25% missing data were removed from consideration. Next, within each subset, PCA plots were again generated for remaining variables to assess their relationship with gene expression. Subsequent differential expression (DE) testing and gene set analyses were performed on data subsets individually using only the covariates appropriate for each subset.

PCA plots of RNA counts demonstrated that after batch correction, tissue site was the primary driver of variation (Supplementary Fig. S1). Therefore, we restricted our analyses to liver, colon, and rectum samples (with liver assessed separately from colon/rectum) given small numbers in other metastatic sites (Table 1). Clinical stage was missing for 37% of patients with RNA-Seq results from the colon/rectum or liver, thus was not considered further. PCA plots were generated and labeled by MSI-status and tumor grade, and tumor tissue site for the colon/rectum subset. Given the small number of patients with MSI-high tumors (Supplementary Table S9) and differences in gene expression by MSI status (Supplementary Fig. S2), MSI-high tumors were excluded from this analysis. There was notable clustering of patients with missing tumor grade for the colon/rectum group with 21% of patients missing grade (Supplementary Fig. S3). Given the presence of strong clustering by grade, and grade likely missing not at random, a missing indicator approach was used. This method has been shown to produce an almost unbiased result while preserving power lost under complete case analysis.^33^ Final variables included for multivariable analysis were tumor grade, gender, early versus average onset, colon vs rectum tumor tissue site (not present for liver subgroup), and either imputed race and ethnicity categories or pivot coordinates for genetic ancestry proportions.

### Gene set analysis workflow 1: GSVA

Because gene set testing approaches test somewhat different hypotheses, we performed gene set analysis in the Hallmark and C2 Biocarta gene sets (342 total) from MSigDB using two distinct workflows.^34–36^ GSVA is a method that evaluates the expression of genes within a gene set relative to those outside of the set (i.e., it is a “competitive” test) and is useful for singling out a few gene sets among many that are associated with a phenotype of interest. On the other hand, mROAST is a method that is focused only on genes within a set (it is “self-contained”) and is more powerful for detecting subtle differences among phenotypes. The first workflow began with filtering the data to retain only genes with at least 10 read counts in greater than 5% of samples, followed by VST^31^ and removeBatchEffects.^32^ These data were then processed by Gene Set Variation Analysis (GSVA) to produce enrichment scores for each sample and gene set.^37^ Differential expression at the gene set level was assessed using a multivariate linear model and the empirical Bayes method in LIMMA. For each data subset, models were run once for the imputed race and ethnicity categories with NH White as the reference group and 5 times for each set of pivot coordinates. P-value correction was performed with the Benjamini-Hochberg method (FDR < 0.05).

### Gene set analysis workflow 2: mROAST

The second gene set analysis workflow began with the previously described prevalence filtering followed by trimmed mean of M values (TMM) normalization and variance modeling at the observational level (VOOM) to generate precision weights.^32^ We then performed gene set testing using the multiple rotation gene set test (mROAST, n rotations = 20,000, mean set statistic, mid-p-values).^38^ RNA assay version was included as a covariate in mROAST and p-values were corrected per Benjamini-Hochberg. For each data subset, mROAST was performed once for each of the imputed race and ethnicity categories with NH White as the reference group for four total tests, and once for each genetic ancestry proportion for five total tests, each on the appropriate set of pivot coordinates.

To maximize robustness of findings, we required an FDR<0.05 in both mROAST and GSVA to report a gene set as significantly enriched in a race and ethnicity imputed category, genetic ancestry proportion, or onset age group.

### Consensus molecular subtypes

Consensus molecular subtypes (CMS) analysis was applied only to samples with colon or rectum as the sequenced tissue site.^39,40^ The CMScaller function assigned each sample a CMS, and a chi-squared test with post-hoc inspection of standardized residuals was used to assess the relationship between CMS and imputed race and ethnicity categories. We further assessed this relationship stratified by age of onset category (EO vs AO). For testing the association of CMS with genetic ancestry proportions, five separate multinomial logistic regressions were performed, each with the five CMS classes as dependent variables and genetic ancestry proportions (as pivot coordinate sets) as the independent variables. Finally, we repeated the multinomial logistic regression stratified by age of onset category.

### Software

Somatic mutation analyses were performed with R version 4.1.3. RNA analyses were performed with R version 4.2.2. RNA-Seq data preparation and analysis steps are diagrammed in Supplementary Fig. S4.

## Results

Patient characteristics are summarized in Table 1. Among the cohort of 8,454 CRC patients, 5,169 (61%) had a matched normal tissue sample and 2,745 (32%) had RNA sequencing performed on the tumor sample. The median age was 60.7 years (IQR 51.1-69.6) with 1,792 (25.6% of patients with available diagnosis age) diagnosed under the age of 50 (i.e., with EOCRC) and 7,997 (94.6%) with microsatellite stable (MSS) disease. Most patients had advanced disease, with 4,254 (81% of those with known stage) diagnosed with stage IV disease. Onset age group differed by imputed race and ethnicity category (p<0.001, Table 1). The Hispanic/Latino category had the highest proportion of EOCRC (37%), while NH White had the lowest proportion (23%). See Supplementary Tables S2 and S3 for patient characteristics stratified by MSI status, and Supplementary Tables S3 and S4 for patient characteristics stratified by onset age group.

### Associations between genetic ancestry and somatic mutations in MSS tumors

Among patients with MSS disease, we examined associations between genetic ancestry proportions and imputed race and ethnicity with protein-altering mutations in 79 genes (Supplementary Table S6, see Methods for selection criteria), somatic copy number alterations (SCNAs) in nine genes, and actionable mutations (present in OncoKB, cf. Methods) in three genes (*BRAF, KRAS, PIK3CA*). Results for MSS tumors are summarized in Table 2 and Figs. 1 and 2.

**Figure 1.**
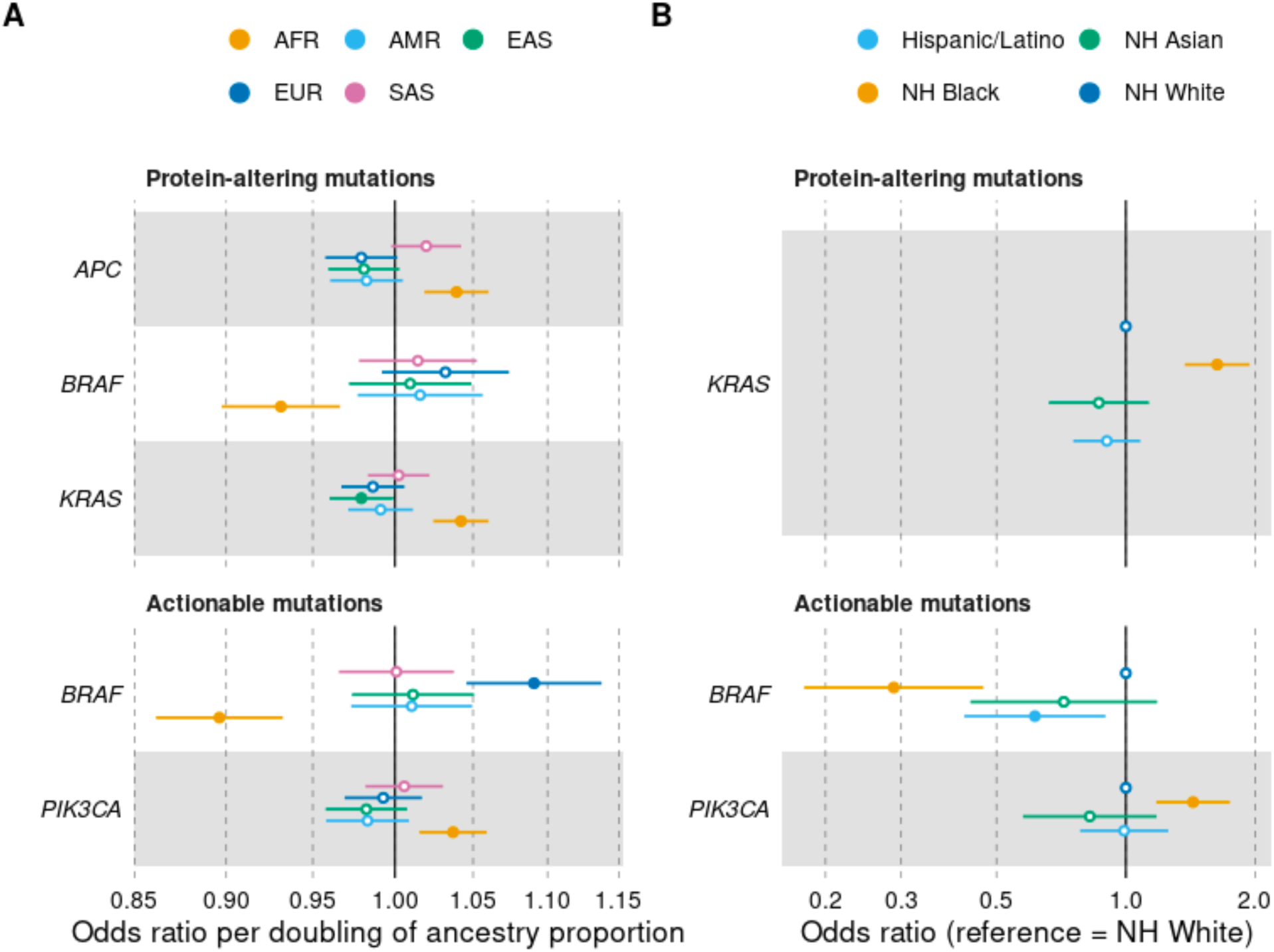
Associations of somatic mutations with genetic ancestry proportions and imputed race and ethnicity categories in patients with MSS disease. **A**, Associations with genetic ancestry proportions. AFR: Africa, AMR: the Americas, EAS: East Asia, EUR: Europe, SAS: South Asia. Odds ratios are with respect to a doubling of a specific genetic ancestry proportion and are adjusted for assay version, gender, age at sample collection, and the other four genetic ancestry proportions. **B**, Associations with imputed race and ethnicity category. Odds ratios are with respect to the NH White race and ethnicity category and are adjusted for assay version, gender, and age at sample collection. Filled circles indicate a logistic regression p<0.05, while open circles indicate p>0.05.

**Figure 2:**
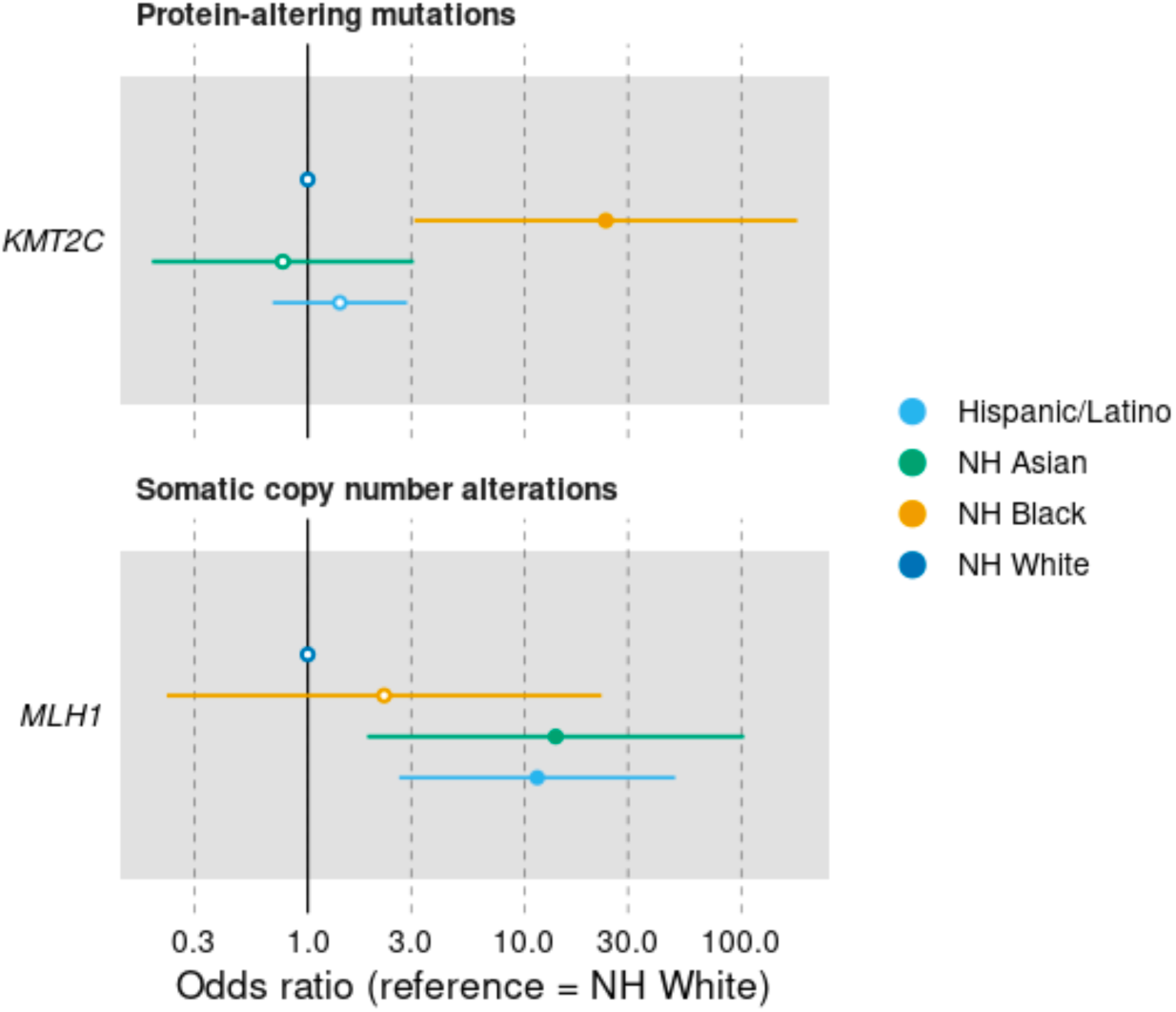
Somatic mutation associations with imputed race and ethnicity categories in patients with MSI-high disease. Odds ratios are with respect to the NH White race and ethnicity category and are adjusted for assay version, gender, and age at sample collection. Filled circles indicate a logistic regression p<0.05, while open circles indicate p>0.05.

**Table 2:** Somatic mutation associations with ancestry proportions and imputed race categories in MSS patients. Mutation type: type of mutation tested. “Actionable” refers to protein-altering mutations that are classified as OncoKB Therapeutic Level of Evidence V2 designation of therapeutic level 1 or 2, or resistance level R1, irrespective of the solid cancer type. Gene: HGNC gene symbol of tested gene. N total: total number of patients included in models. N with mutation = number of patients included in models who have one or more of the mutation type in the gene. *P* LR (FDR): *P*-value for likelihood ratio test, adjusted to control the false discovery rate. Ancestry or imputed race group: ancestry or imputed race group associated with the presence/absence of mutations in this gene in logistic regression test. OR (95% CI): odds ratio per doubling of genetic ancestry proportion (in the case of ancestry) or odds ratio compared to NH White category (in the case of imputed race group) and 95% confidence interval in the logistic regression test. *P* Logistic = *P*-value for the specific ancestry proportion or imputed race group in the logistic regression test, not adjusted for multiple tests.

| Mutation type | Gene | N total | N with mutation | *P* LR (FDR) | Ancestry or imputed race/ ethnicity group | OR (95% CI) | *P* Logistic |
| --- | --- | --- | --- | --- | --- | --- | --- |
| Protein-altering | *APC* | 4,871 | 3,558 | 0.007 | AFR | 1.04 (1.02, 1.06) | 1.6e-04 |
| Protein-altering | *BRAF* | 4,871 | 365 | 0.047 | AFR | 0.93 (0.90, 0.97) | 1.6e-04 |
| Protein-altering | *KRAS* | 4,871 | 2,343 | 0.004 | AFR | 1.04 (1.02, 1.06) | 2.9e-06 |
|  |  |  |  |  | EAS | 0.98 (0.96, 0.999) | 0.039 |
| Actionable | *BRAF* | 7,965 | 402 | <1e-08 | AFR | 0.90 (0.86, 0.93) | 5.7e-08 |
|  |  |  |  |  | EUR | 1.09 (1.06, 1.14) | 5.4e-05 |
| Actionable | *PIK3CA* | 7,965 | 855 | 0.016 | AFR | 1.04 (1.02, 1.06) | 7.1e-04 |
| Protein-altering | *KRAS* | 4,723 | 2,276 | 3.2e-06 | NH Black | 1.63 (1.37, 1.94) | 3.3e-08 |
| Actionable | *BRAF* | 7,718 | 388 | 1.8e-08 | Hispanic/<br>Latino | 0.61 (0.42, 0.90) | 0.012 |
|  |  |  |  |  | NH Black | 0.29 (0.18, 0.47) | 3.9e-07 |
| Actionable | *PIK3CA* | 7,718 | 831 | 0.003 | NH Black | 1.43 (1.18, 1.75) | 3.4e-04 |

Increased AFR ancestry was associated with higher odds of protein-altering mutations (Fig. 1A) in *APC* [odds ratio (OR) per doubling of ancestry proportion, 1.04; 95% confidence interval (CI), 1.02-1.06] and *KRAS* (OR, 1.04; 95% CI, 1.02-1.06), along with decreased odds of such mutations in *BRAF* (OR, 0.93; 95% CI, 0.90-0.97). EAS genetic ancestry was associated with decreased odds of protein-altering mutations in *KRAS* (OR, 0.98; 95% CI, 0.96-0.999). For actionable mutations (Fig. 1A), increased AFR genetic ancestry was associated with increased odds of *PIK3CA* mutations (OR, 1.04; 95% CI 1.02-1.06), and decreased odds of *BRAF* mutations (OR, 0.90; 95% CI, 0.86-0.93). Increased EUR genetic ancestry was positively associated with actionable mutations in *BRAF* (OR, 1.09; 95% CI, 1.06-1.14). No genetic ancestry proportion associations were found with SCNAs.

In tests of imputed race and ethnicity categories, we found that NH Blacks had higher odds of protein-altering mutations (Fig. 1B) in *KRAS* compared to NH Whites (OR, 1.63, 95% CI 1.37-1.94). The association was not significant for actionable mutations (Fig. 1B). NH Black and Hispanic/Latino patients had lower odds of actionable mutations of *BRAF* (OR, 0.61; 95% CI 0.42-0.90 and OR, 0.29; 95% CI, 0.18-0.47, respectively) compared to NH White patients, while NH Blacks had higher odds of actionable mutations in *PIK3CA* (OR, 1.43; 95% CI, 1.18-1.75).

### Somatic mutation associations with interaction effects in MSS tumors

Two genes showed different imputed race and ethnicity category associations by either diagnosis age or primary site. Hispanic/Latino patients with AOCRC had higher odds of *FLT3* SCNAs than NH White AOCRC patients (OR, 2.38; 95% CI, 1.53-3.72), while no association was present in those with EOCRC (OR, 0.46; 95% CI 0.18-1.18; Fig. 3, Supplementary Table S7). Both NH Asian and NH Black patients with primary tumors in the colon showed decreased odds of actionable mutations in *BRAF* compared to NH Whites (OR, 0.12; 95% CI, 0.16-0.86 and OR, 0.10; 95% CI, 0.03-0.42, respectively), with no association seen for rectal tumors (OR, 1.84; 95% CI, 0.41-8.32 and OR, 2.12; 95% CI, 0.68-6.59, respectively (Figure 3, Supplementary Table S7).

**Figure 3:**
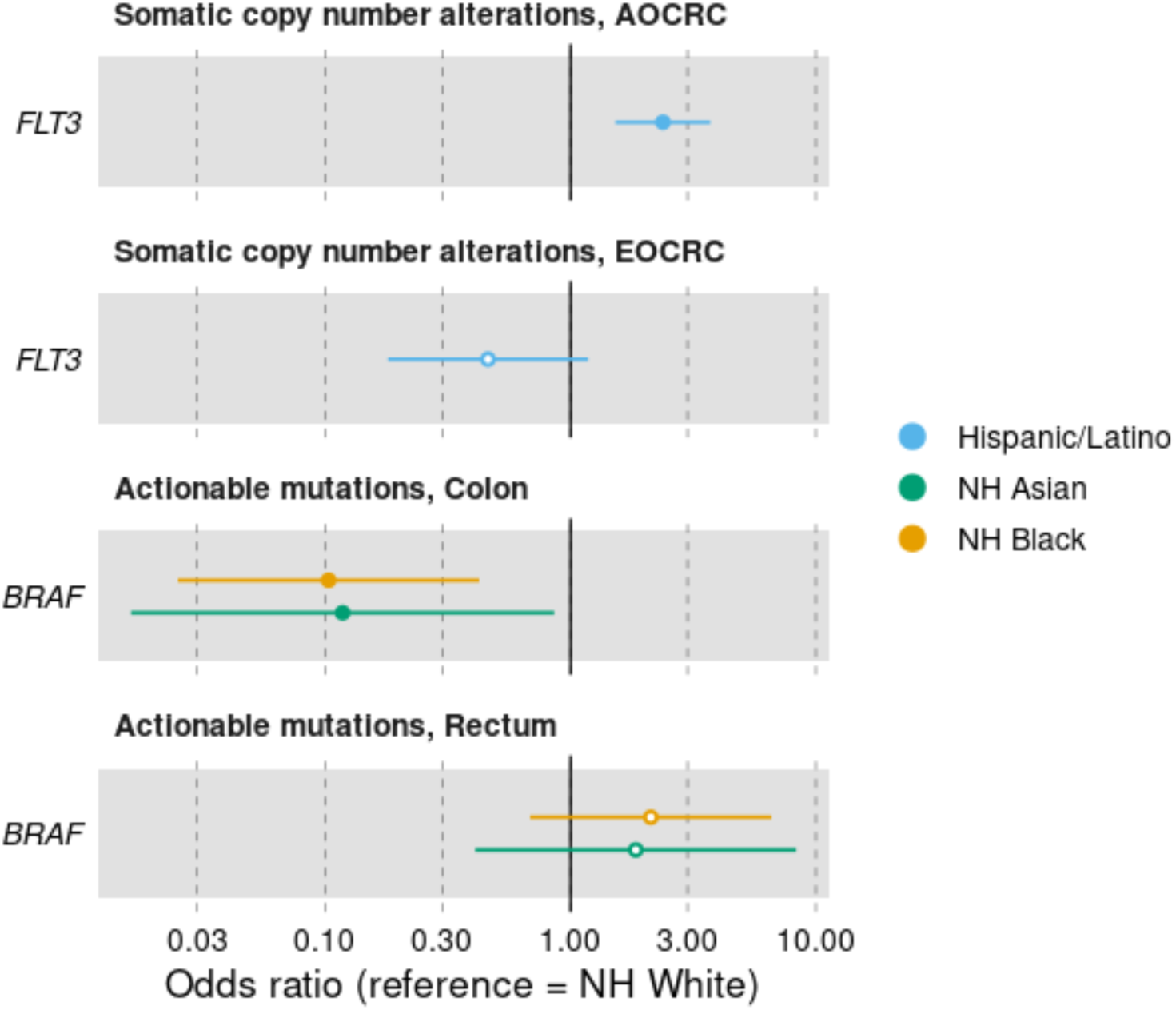
Interaction effects by onset age group or by primary tumor site in somatic mutation associations with imputed race and ethnicity categories in patients with MSS disease. Odds ratios are with respect to the NH White race and ethnicity category and are adjusted for assay version, gender, and age at sample collection. Filled circles indicate a logistic regression p<0.05, while open circles indicate p>0.05.

### Somatic mutation associations in MSI-high tumors

Approximately 5% of the cohort had MSI-high tumors, which ranged from 3.6% to 5.9% in patients with imputed NH Asian and Hispanic/Latino race and ethnicity, respectively (Table 1). The difference in proportion of MSI-high tumors by race and ethnicity category was not significant (p=0.062). Among patients with AOCRC, prevalence of MSI-high tumors differed by imputed race and ethnicity (p=0.008, Supplementary Table S4). Hispanic/Latino and NH White patients had the highest proportion of MSI-high tumors at 6.0% and 6.4%, respectively. In contrast, in EOCRC, patients with NH Black (5.4%), Hispanic/Latino (6.1%) and complex (6.9%) imputed race and ethnicity had higher rates of MSI-high tumors (Supplementary Table S5), though the differences were not statistically significant (p=0.074). Among MSI-high patients, we tested the association of genetic ancestry proportions and imputed race and ethnicity with the presence of protein-altering mutations in 127 genes, SCNAs in two genes, and actionable mutations in two genes (Supplementary Table S4). No associations were found between genetic ancestry proportions and the presence of any mutations. NH Black MSI-high patients had higher odds of having protein-altering mutations in *KMT2C* compared to NH Whites (OR, 23.7; 95% CI, 3.1-181; Figure 2A, Supplementary Table S8), and NH Asian and Hispanic/Latino MSI-high patients were more likely to have *MLH1* SCNAs compared to NH Whites (OR, 13.9; 95% CI, 1.9-103 and OR, 11.4; 95% CI, 2.6-49.6, respectively; Figure 2B, Supplementary Table S8).

### TMB in MSI-high tumors

Among MSI-high patients, there was no statistically significant difference of TMB by imputed race and ethnicity group (p=0.21), or onset group (p=0.85), nor was TMB significantly different among the subset of MSI-high patients with AOCRC (p=0.06) or EOCRC (p=0.26); see Fig. 4A-D.

**Figure 4.**
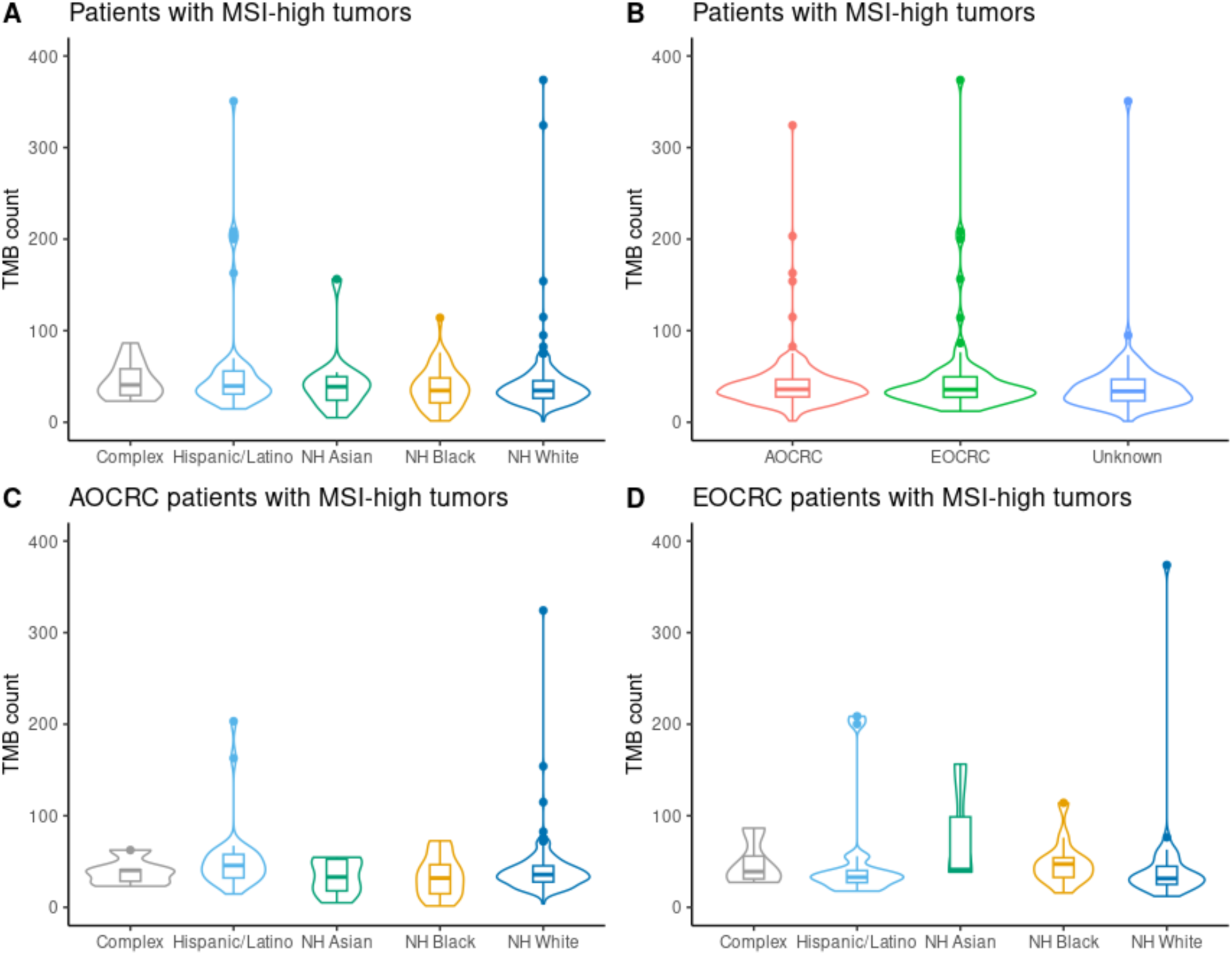
Distribution of TMB count by imputed race and ethnicity group and age of onset for MSI-high patients. **A**, TMB count by imputed race and ethnicity group. **B**, TMB count by age of onset. **C**, TMB count in AOCRC patients by race and ethnicity group. **D**, TMB count in EOCRC patients by race and ethnicity group.

### Variable selection for mRNA analyses

PCA plots of RNA counts demonstrated that after batch correction, tissue site was the primary driver of variation (Supplementary Fig. S1). Therefore, we restricted our analyses to liver, colon, and rectum samples (with liver assessed separately from colon/rectum) given small numbers in other metastatic sites (Table 1). Clinical stage was missing for 378% of patients with RNA-Seq results from the colon/rectum or liver, thus was not considered further. PCA plots were generated and labeled by MSI-status and tumor grade, and tumor tissue site for the colon/rectum subset. Given the small number of MSI-high patients (Supplementary Table S9) and differences in gene expression by MSI status (Supplementary Fig. S2), MSI-high patients were excluded from this analysis. There was notable clustering by missing tumor grade in both PCA and UMAP for the colon/rectum group with 21% of patients missing grade (Supplementary Fig. S3). Given the presence of strong clustering by grade, and grade likely missing not at random, a missing indicator approach was used. This method has been shown to produce an almost unbiased result while preserving power lost under complete case analysis.^33^ Final variables included for multivariable analysis were tumor grade, gender, early versus average onset, colon vs rectum tumor tissue site (not present for liver subgroup), and either imputed race and ethnicity categories or pivot coordinates for genetic ancestry proportions.

### Associations between genetic ancestry and expression of gene sets

We next examined associations between genetic ancestry or imputed race and ethnicity category with expression of genes in the Hallmark and Biocarta C2 gene sets (342 total). In MSS colon/rectum samples (n=1,830), the imputed NH Black category was consistently associated with under-expression compared to the NH White category in the following gene sets: Hallmark coagulation (mROAST p=0.021, GSVA p=0.005), BioCarta alternative complement (mROAST p=0.009, GSVA p=0.005), BioCarta RECK (mROAST p=0.026, GSVA p=0.007), and BioCarta Rhodopsin (mROAST p=0.026, GSVA p=0.038; Table 3). Highly differentially expressed genes in these gene sets included: complement factor *C3*, tissue inhibitors of metalloproteinases *TIMP2* and *TIMP3*, matrix metallopeptidase 11 (*MMP11*), coagulation factor VIII (*F8)*, cathepsin K (*CTSK)*, and antithrombin III (*SERPINC1)* (Supplementary Table S10.1). Significant underexpression associated with increased AFR genetic ancestry in the above gene sets was found only by GSVA; we include the AFR results in Table 3 for comparison.

**Table 3:** RNA gene set results. Gene sets are reported as significant only if the corrected p value from both mROAST and GSVA was < 0.05 and at least 50% of individual genes in the set were significantly differentially expressed as reported by mROAST. mROAST uses more conservative mid-p values during FDR correction. Included in this table alongside significant results is the result for the genetic ancestry proportion (or imputed group) that has the most overlap with the significant finding. All results reported in this table for the Non-Hispanic Black imputed category are underexpression of the gene set in comparison to Non-Hispanic White; all results for AFR represent decreased gene expression as the dominance of AFR compared to other ancestries increases.

| Gene Set | Genetic Ancestry | Tissue Site | mROAST Proportion of Genes in Set Significantly Underexpressed | mROAST FDR | GSVA FDR |
| --- | --- | --- | --- | --- | --- |
| Hallmark Coagulation |  |  |  |  |  |
|  | NH Black | Colon/Rectum | 65/130 | 0.021 | 0.005 |
|  | AFR | Colon/Rectum | 60/130 | 0.083 | 0.038 |
| BioCarta Alternative Complement |  |  |  |  |  |
|  | NH Black | Colon/Rectum | 5/8 | 0.009 | 0.005 |
|  | AFR | Colon/Rectum | 3/8 | 0.083 | 0.046 |
| BioCarta RECK |  |  |  |  |  |
|  | NH Black | Colon/Rectum | 7/9 | 0.026 | 0.007 |
|  | AFR | Colon/Rectum | 7/9 | 0.105 | 0.042 |
| BioCarta Rhodopsin |  |  |  |  |  |
|  | NH Black | Colon/Rectum | 5/6 | 0.026 | 0.011 |
|  | AFR | Colon/Rectum | 5/6 | 0.077 | 0.038 |
| CREM |  |  |  |  |  |
|  | NH Black | Liver | 3/5 | 0.528 | 0.079 |
|  | AFR | Liver | 3/5 | 0.026 | 0.002 |

In MSS liver samples (n=778), greater AFR genetic ancestry (but not the NH Black imputed category) was associated with underexpression in the BioCarta CREM gene set (Table 3 and Supplementary Table S10.2). There were no significant findings by age of onset group.

### Associations between genetic ancestry and CRC consensus molecular subtypes (CMS)

Among 1,957 patients where CMS were obtained with CMScaller,^40^ including both MSS and MSI-H patients, 252 were imputed non-Hispanic (NH) Black, 98 NH Asian, 287 Hispanic/Latino, 66 Complex, and 1,254 NH White. CMS was associated with race and ethnicity imputed categories (p=0.004). Inspection of the standardized chi-square residuals revealed greater than expected NH Black CMS3 (66 observed vs 46 expected, p=0.001), less than expected NH Black CMS1 (18 vs 30, p=0.011), and greater than expected Hispanic/Latino indeterminate CMS (36 vs 26, p=0.031), (Figure 5, Supplementary Table S12). When stratifying by age of onset group the overall chi-square test of independence was no longer significant for EOCRC but remained significant for AOCRC, and inspection of standardized residuals revealed the association of indeterminate CMS and Hispanic/Latino imputed category was only present among EO, while the association of NH Black and CMS1 and CMS3 was only present among AO.

**Figure 5:**
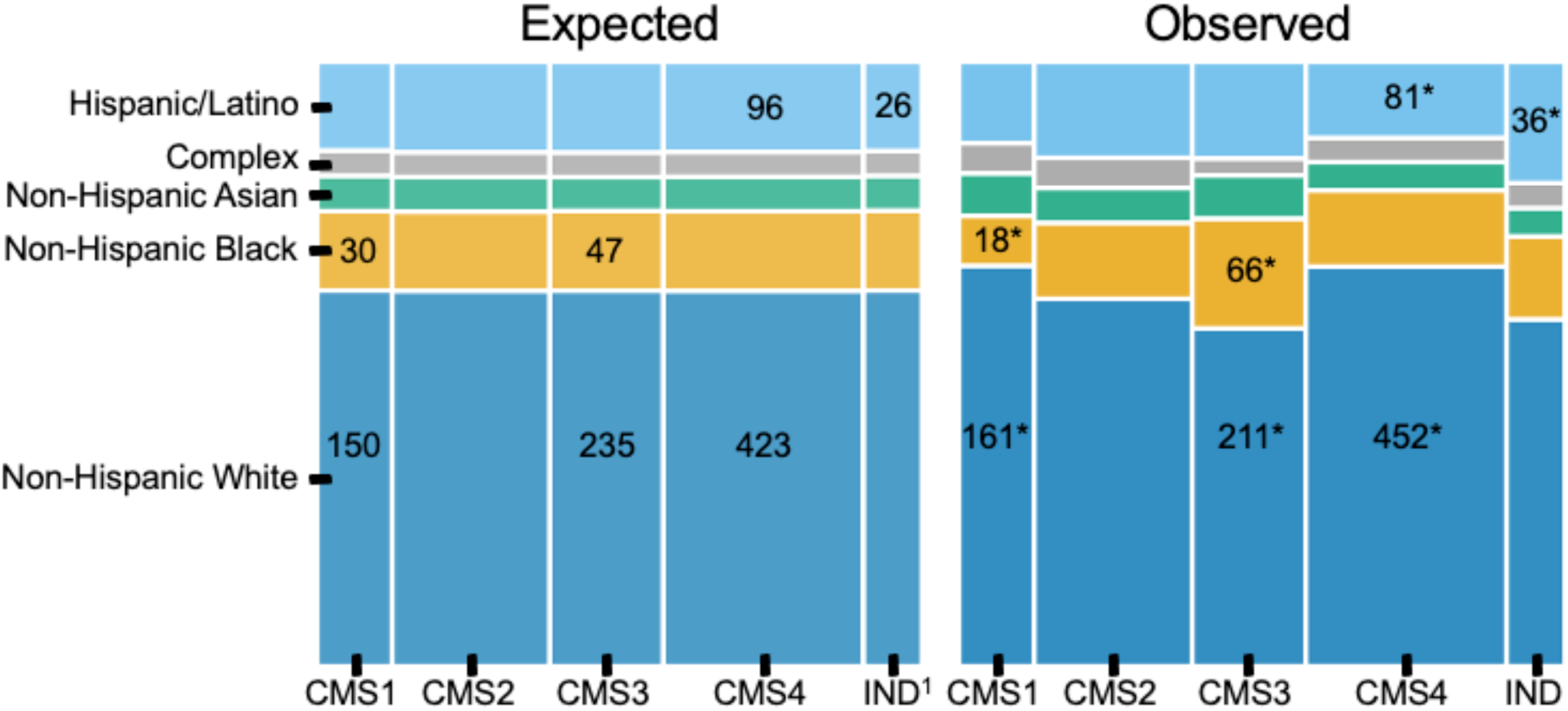
Expected vs observed proportions of patients by imputed racial/ethnic category and consensus molecular subtype. Panel area in the expected block is proportional to the null hypothesis of equal racial and ethnic distribution across CMS. The imputed category bar height is equal to the proportion of that category in the cohort population and under the null hypothesis is constant across CMS. Panel area in the observed block is proportional to the observed racial and ethnic distribution across CMS. Bar width for each CMS is proportional to the observed patient assignments. Findings with absolute standardized residuals >1.96 indicated with * (chi-square test of independence). ^1^Indeterminate CMS

In the analysis of genetic ancestry proportions, increased AFR genetic ancestry was significantly associated with CMS3 (OR, 1.056 per doubling of AFR proportion, 95% CI, 1.003-1.111) and indeterminate CMS (OR, 1.083; 95% CI, 1.021-1.149), while increased EUR genetic ancestry was associated with decreased odds of CMS3 (OR, 0.925; 95% CI, 0.874-0.979), all with CMS1 as the reference outcome (Supplementary Table S13). When stratifying by age of onset group these associations were only statistically significant in the AOCRC group.

## Discussion

Using comprehensive tumor profiling in a large, diverse patient cohort, we found differences in somatic mutation and gene expression by genetic ancestry in CRC. Prior studies have used self-reported or observed race and ethnicity or have used genetic ancestry to categorize individuals into groups, excluding individuals who do not meet selected thresholds. Instead, our study uses genetic ancestry proportions directly to look for associations, using methods that control for correlations among ancestries. Further, we leverage genetic ancestry to impute race and ethnicity categories to address missingness in real world data.

Given rising incidence of EOCRC, we first sought to assess differences in imputed race and ethnicity with tumor genetic profile by age of onset. However, in our data, no significant interactions were found except for *FLT3,* which had higher odds of SCNAs in Hispanic/Latino patients with AOCRC but not EOCRC.

For MSS CRC across all ages, NH Black patients and those with greater AFR genetic ancestry had lower odds of actionable variants in *BRAF* and higher odds of short protein-altering mutations in *KRAS*. In contrast, increased EAS ancestry was associated with decreased odds of protein-altering *KRAS* mutations and greater odds of actionable *BRAF* mutations. *KRAS*-WT and left-sided tumors are approved for treatment with *EGFR* inhibitors such as cetuximab.^41^ Our findings are consistent with previously published studies using self-reported race showing higher rates of *KRAS* mutations in NH Black patients,^10,12,42^ underscoring the importance of assessing targeted therapies in diverse populations. Standard of care for patients with metastatic cancer harboring mutations in BRAF V600E include combination BRAF and EGFR inhibitors.^43^ These mutations usually portend a poor prognosis with early development of metastatic disease and were less common in patients with imputed NH Black and Hispanic/Latino race and ethnicity and more common in patients with increased EUR ancestry.^12^

Approximately 20-25% of CRC patients harbor activating mutations in *PIK3CA*, which activates the mTOR pathway.^44^ Some studies have suggested that the presence of *PIK3CA* mutations could confer resistance to first-line chemotherapy, although the data are preliminary.^45^ Inhibitors of PIK3CA have been approved for PIK3CA positive treatment resistant metastatic breast cancer.^46^ As such, PIK3CA mutations could represent an opportunity for targeted therapy in CRC, particularly in combination with other drugs, since *PIK3CA* mutations are also associated with higher rates of mutations in genes in other key cancer pathways.^44^ The higher rate of actionable *PIK3CA* mutations in MSS patients with greater AFR ancestry and imputed NH Black race suggests these combinations could preferentially benefit minority subgroups with CRC.

Thrombosis is one of the leading causes of death among cancer patients,^47^ and there is increased risk of both overall and cancer associated thrombosis among Black patients.^48,49^ In our study, the Hallmark coagulation gene set was significantly underexpressed in tumors from NH Black patients. Specifically, coagulation factors *F7* and *F11* and platelet tissue factor *TF* were under-expressed, while antithrombin III *SERPINC1* was overexpressed in NH Black patients compared to NH White. These findings do not support that changes in tumor coagulation gene expression pathways contribute to the elevated thrombosis risk observed in Black CRC patients.

Patients with imputed NH Black race and ethnicity or increased AFR ancestry had higher odds of CMS3 tumors. So called “metabolic tumors,” CMS3 tumors display marked metabolic deregulation with the majority harboring mutations in *KRAS*.^39^ As such, this finding is concordant with the positive association of *KRAS* mutations and AFR ancestry found in our study. Hispanic/Latino patients were assigned to the indeterminate CMS category more often than expected. While the reason for this is unclear, one possibility is the underrepresentation of non-White patients in the datasets used to define CMS.^23,39^ As future trials and drug development efforts may stratify patients by CMS, it is important to ensure these categorizations accurately represent a diverse CRC patient population.

Limitations of our study include incomplete metadata on clinical stage, sidedness, age of diagnosis, tumor grade, and normal tissue sequencing. Also, we were not able to impute “American Indian or Alaska Native” or “Native Hawaiian or Other Pacific Islander” categories due to limitations in the public reference allele frequencies, and the small number of patients of such categories in our cohort (estimated <1%). These patients may be misclassified as Hispanic/Latino or NH Asian, respectively (cf. Table 1), however given the small number we do not expect these to significantly change our findings regarding imputed Hispanic/Latino or NH Asian categories.

The strengths of our study include large sample size and the use of genetic ancestry rather than reliance on self-reported or stated race and ethnicity. Our cohort also includes a large fraction of patients for whom matched tumor-normal sequencing data is available, allowing better discrimination between germline variants and somatic mutations. Another strength of our study is the concurrent analysis of genomic somatic mutations with transcriptional profiles of the patient’s tumors. Methodologically, by applying compositional analysis in our logistic regressions, we were able to minimize comparisons involving a single reference group (typically Whites) while controlling for correlations among genetic ancestries when they are reported as proportions that sum to one. Further, we used two distinct gene set analysis and RNAseq normalization methods to demonstrate consistency and strengthen our gene expression findings.

## Conclusions

In summary, through analyzing a large, diverse CRC patient cohort, we found associations between genetic ancestry and prevalence of somatic mutations in CRC driver genes, levels of gene expression in cancer related gene sets, and distribution of consensus molecular subtypes that have not previously been reported in studies using race and ethnicity categories alone. Increased AFR genetic ancestry was associated with higher odds of *APC, KRAS,* and *PIK3CA* mutations and CMS3 tumors, and lower odds of *BRAF* mutations. Increased EAS genetic ancestry correlated with lower odds of mutations in *KRAS*. Furthermore, the increased odds of indeterminate CMS tumors in the imputed Hispanic/Latino category suggests that more diverse representation could reduce disparities in the applicability of disease subtype models. Our findings demonstrate the advantage of using genetic ancestry in studies of disparities in CRC, and highlight the need to validate proposed therapies, biomarkers, and prognosis indicators in diverse patient populations.

## Data Availability

All data supporting the conclusions of this article are included within the article and its additional files. Due to patient privacy and HIPAA regulations our raw data cannot be publicly available.

## Availability of data and materials

The datasets supporting the conclusions of this article are included within the article and its additional files. Due to patient privacy, data sharing consent limitations, and HIPAA regulations, our raw data cannot be submitted to publicly available databases.

## Abbreviations

CRC: Colorectal cancer
EOCRC: Early onset CRC
AOCRC: Average onset CRC
CMS: CRC consensus molecular subtypes
AFR: African continental genetic ancestry
AIM: Ancestry-informative marker
AMR: Americas (Native American) continental genetic ancestry
EAS: East Asian continental genetic ancestry
EUR: European continental genetic ancestry
SAS: South Asian continental genetic ancestry
NH: Non-Hispanic
MSI: Microsatellite instability
MSS: Microsatellite stable disease
SCNA: Somatic copy-number alterations
TMB: Tumor mutational burden
CI: 95% confidence interval
DE: Differential expression
GSVA: Gene-set variation analysis
ILR: Isometric log ratio
LRT: Likelihood ratio test
OR: Odds ratio
PCA: Principal component analysis
TMM: Trimmed median of M values
VST: Variant stabilizing transform
RNA: Ribonucleic acid
RNAseq: RNA sequencing

## Acknowledgements

We would like to thank Yan Liu (Tempus), Carlos D. Bustamante and Alex Ioannidis (Stanford University) for statistical and methodology discussions and advice. We also acknowledge Frank Nothaft, Rafael Esteller, Nick Rigan, and Arvind Prasad from the Tempus Lens team for their superb assistance in procuring de-identified data and correcting data problems needed for this work. We thank Vanessa Nepomuceno from the Tempus Publications team for copyediting the manuscript.

## Autor Information

### Authors and Affiliations

*Tempus Inc, Chicago, IL, USA*

Brooke Rhead, Yannick Pouliot, Justin Guinney, and Francisco M. De La Vega

*Department of Radiation Oncology, University of Texas Southwestern, Dallas, TX 75390, USA*

David M. Hein and Nina N. Sanford

### Corresponding authors

Correspondence to Nina N. Sanford and Francisco M. De La Vega.

### Co-first authors

Brooke Rhead and David M. Hein

### Contributions

Brooke Rhead: Methodology, data analysis, visualization, writing - review, editing. David Hein: Methodology, data analysis, visualization, writing - review, editing. Yannick Pouliot: Data procurement, curation, analysis, methodology, writing - review, editing. Justin Guinney: Methodology, writing - review and editing. Francisco De La Vega: Conceptualization, resources, supervision, writing - review, editing. Nina Sanford: Conceptualization, resources, supervision, writing - original draft, writing - review, editing. All authors reviewed and suggested edits for the final version of the manuscript. The authors read and approved the final manuscript. 34

## Ethics declarations

### Ethics approval and consent to participate

All analyses were performed using de-identified data; The need for Institutional Review Board Approval was exempted by the IRB of Advarrra, Inc., protocol no: Pro00042950, on April 15, 2020.

### Consent for publication

Not applicable.

---

**Supplementary Table S1.** Race and ethnicity imputation from continental genetic ancestry. Mutually-exclusive race ethnicity categories are imputed based on the continental ancestry thresholds indicated in the table, which are based on literature and our own assessment of patient data. Patients who did not fit any of these categories were termed “complex” and included patients with complex continental admixture. We evaluated sensitivity and specificity on 20,396 patients from the Tempus database for whom both stated (self-reported or observed) race and ethnicity were available, except in the case of Hispanic/Latino ethnicity patients for which we did not require a stated race. We created mutually exclusive classes for evaluation by including patients who had a stated race and non-Hispanic ethnicity. Hispanic/Latino patients included any patient with that stated ethnicity, regardless of race metadata. The overall accuracy (% correctly categorized) was 95.4% (ignoring complex).

| Imputed category | Genetic ancestry thresholds | Sensitivity | Specificity |
| --- | --- | --- | --- |
| Non-Hispanic Asian | >70% combined EAS and SAS | 0.922 | 0.999 |
| Non-Hispanic Black | >20% AFR, <10% AMR, and >70% combined AFR and EUR | 0.934 | 0.997 |
| Non-Hispanic White | >80% EUR and <10% AMR | 0.980 | 0.997 |
| Hispanic/Latino | >10% AMR and >70% combined AMR, EUR, and AFR | 0.854 | 0.997 |
| Complex | Remaining patients not meeting above thresholds | N/A | N/A |

**Supplementary Table S2:**
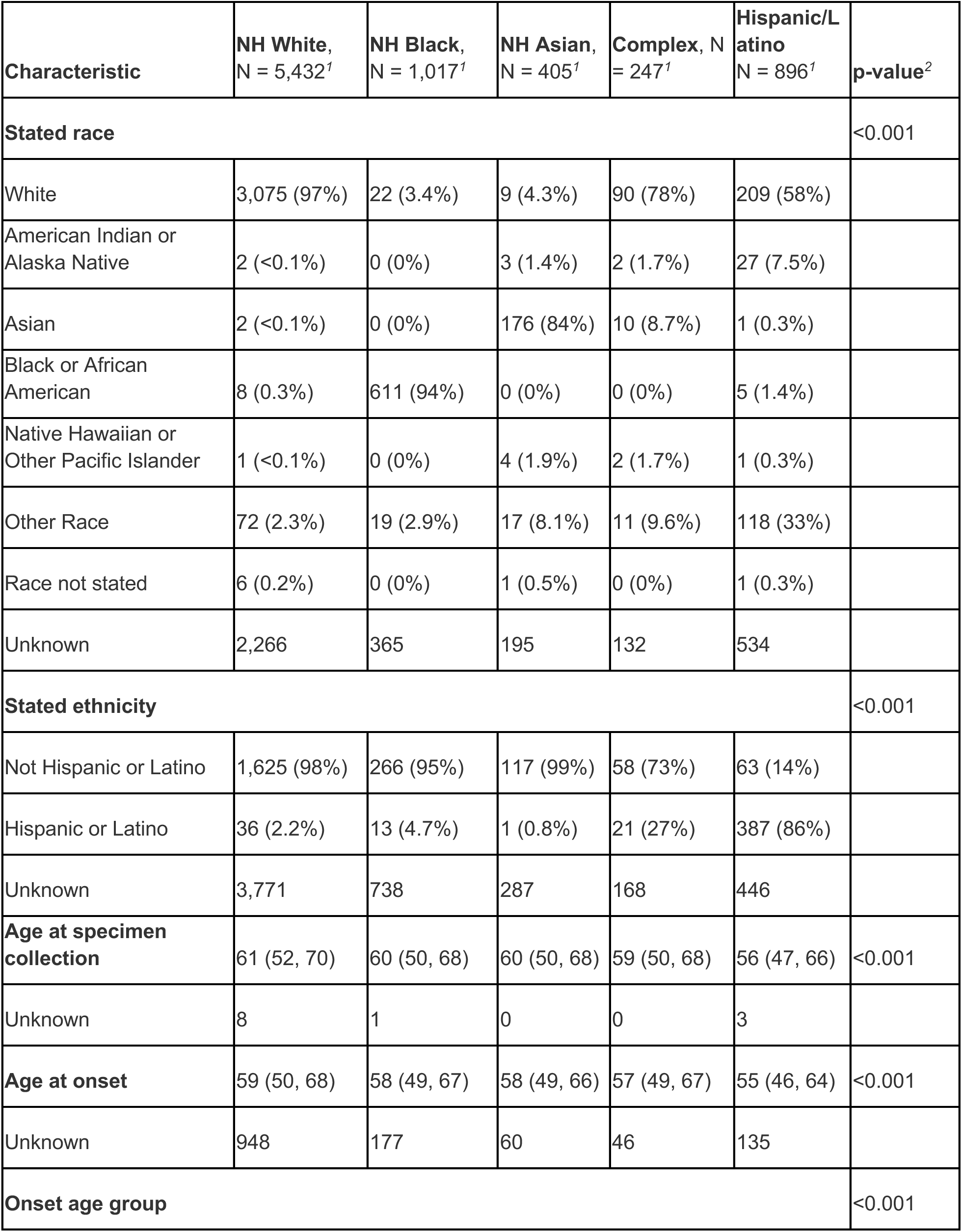
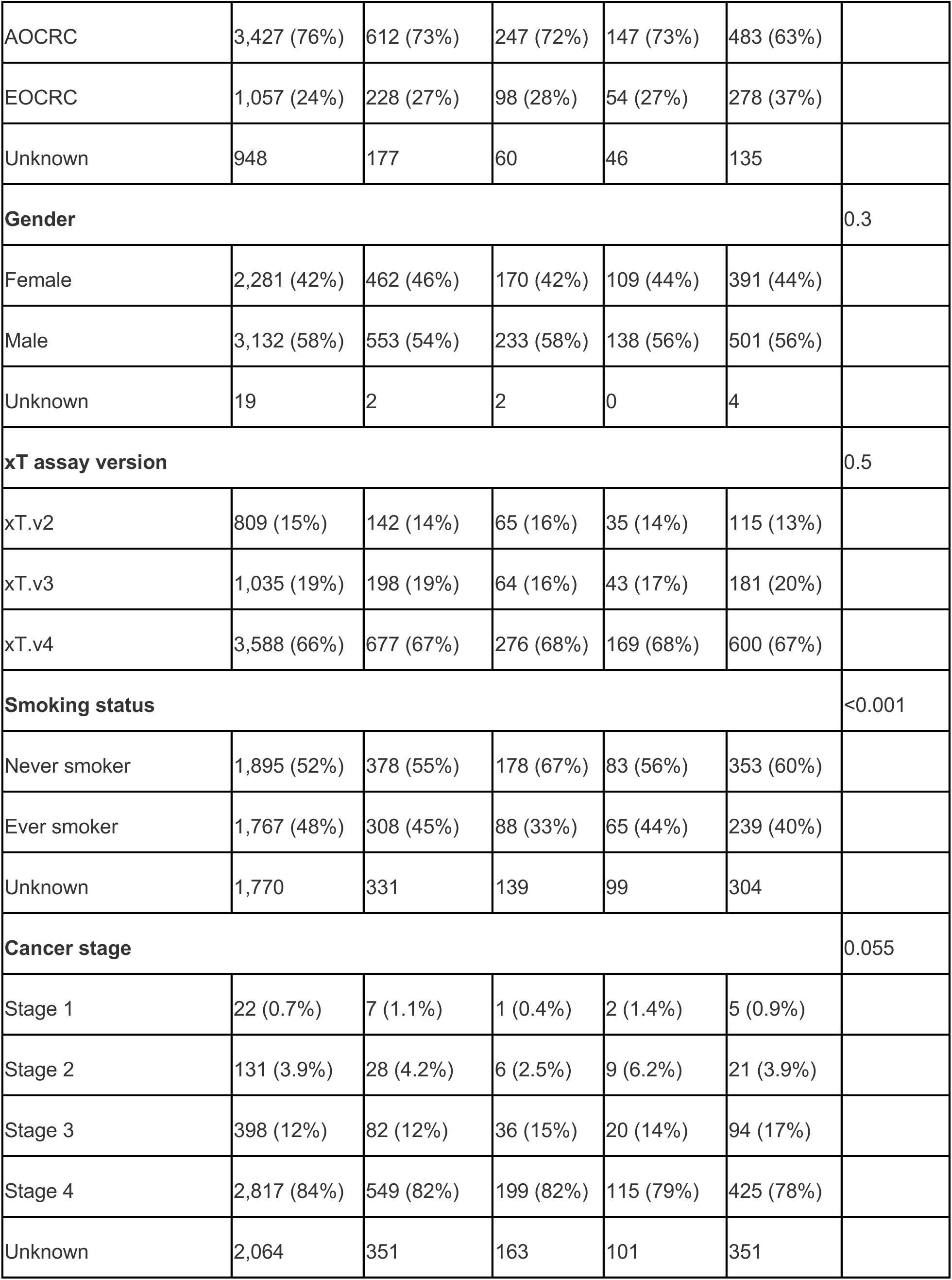
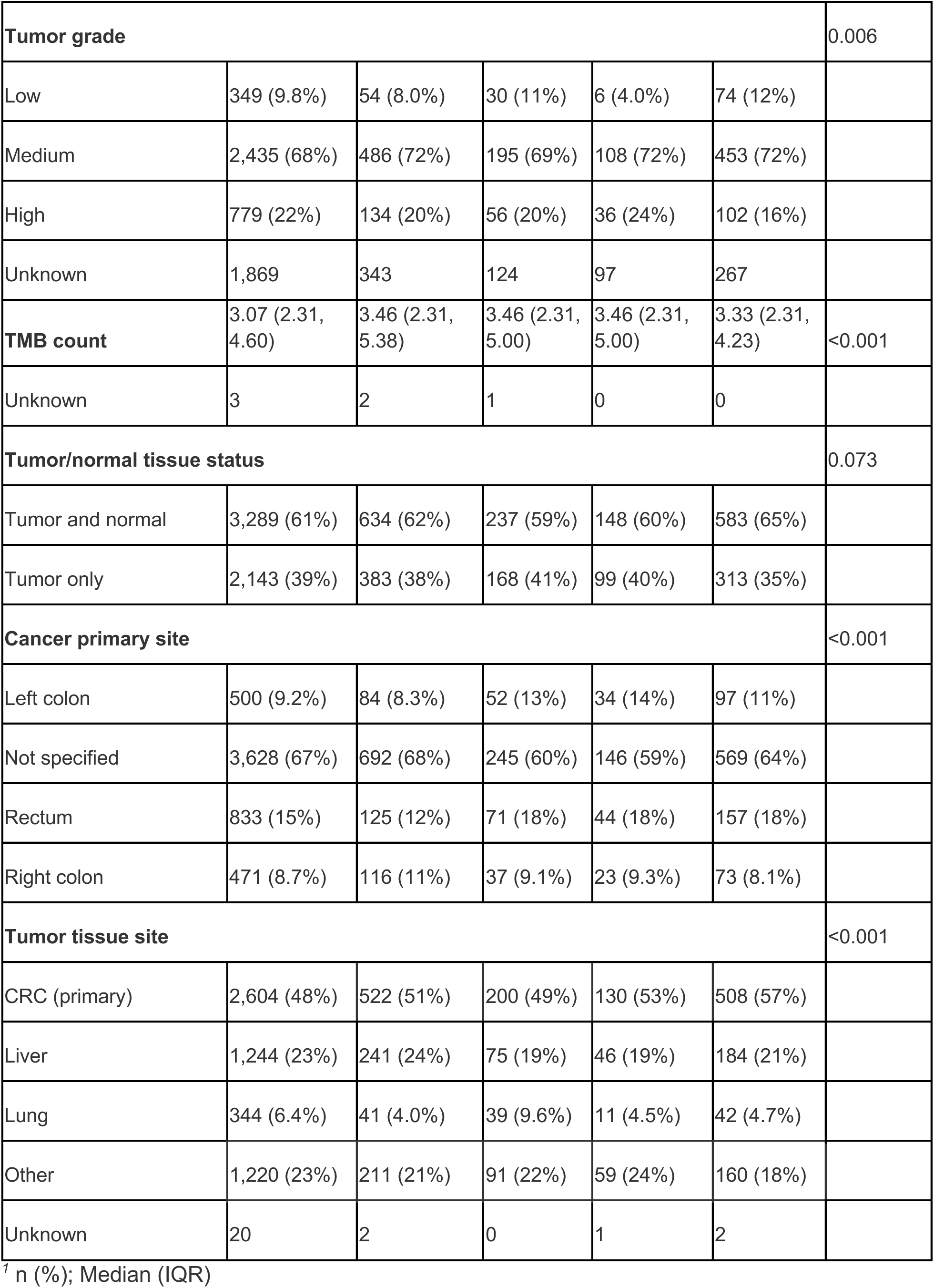
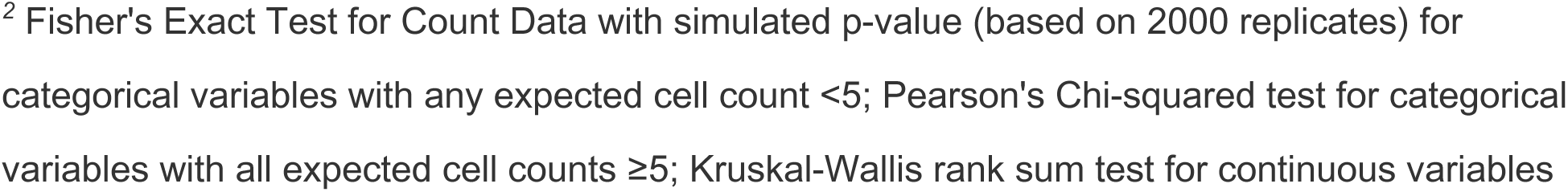
Patient characteristics for MSS patients by imputed race ethnicity category.

**Supplementary Table S3:**
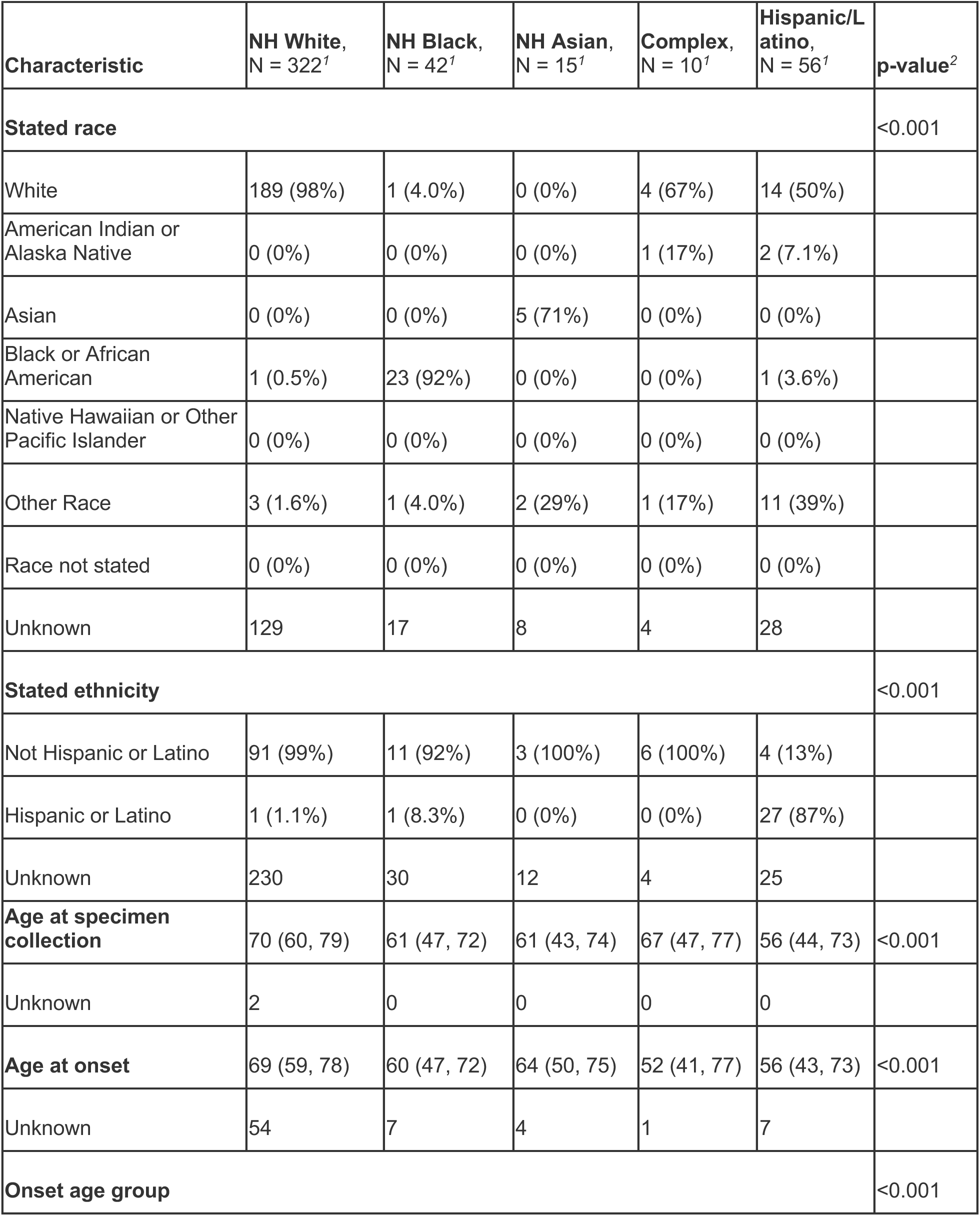
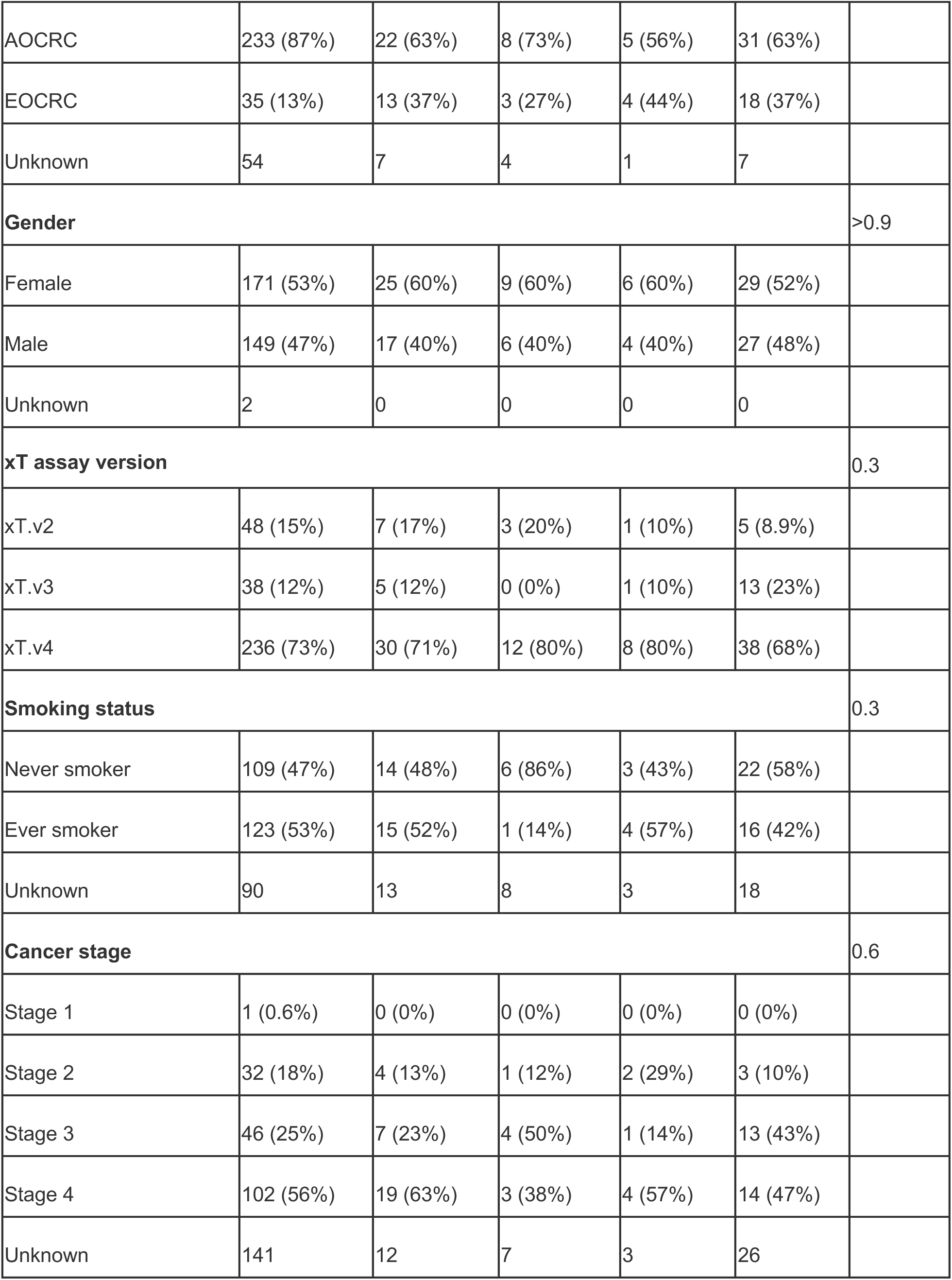
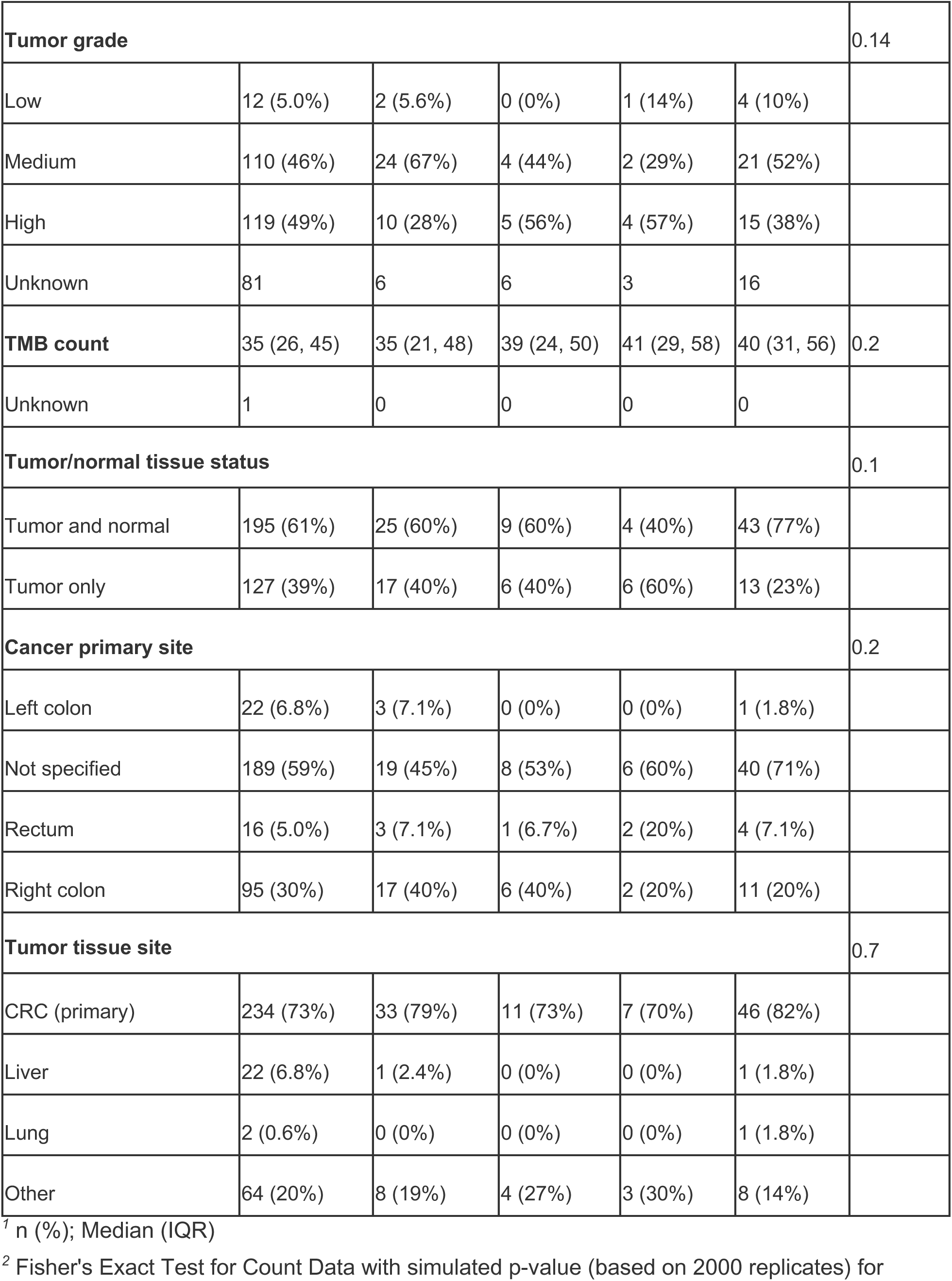
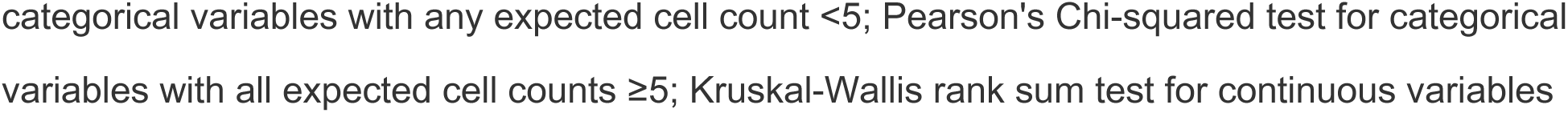
Patient characteristics for MSI-high patients by imputed race ethnicity group.

**Supplementary Table S4.** AOCRC patient characteristics by imputed race and ethnicity group.

| Characteristic | NH White,<br>N = 3,664<sup>1</sup> | NH Black,<br>N = 635<sup>1</sup> | NH Asian,<br>N = 255<sup>1</sup> | Complex,<br>N = 153<sup>1</sup> | Hispanic/L<br>atino,<br>N = 514<sup>1</sup> | p-value<sup>2</sup> |
| --- | --- | --- | --- | --- | --- | --- |
| **Stated race** |  |  |  |  |  | <0.001 |
| White | 2,117 (98%) | 13 (3.2%) | 6 (4.4%) | 62 (84%) | 131 (58%) |  |
| American Indian or<br>Alaska Native | 1 (<0.1%) | 0 (0%) | 2 (1.5%) | 1 (1.4%) | 16 (7.1%) |  |
| Asian | 2 (<0.1%) | 0 (0%) | 113 (84%) | 5 (6.8%) | 1 (0.4%) |  |
| Black or African<br>American | 3 (0.1%) | 385 (95%) | 0 (0%) | 0 (0%) | 3 (1.3%) |  |
| Native Hawaiian or<br>Other Pacific Islander | 1 (<0.1%) | 0 (0%) | 2 (1.5%) | 1 (1.4%) | 0 (0%) |  |
| Other Race | 44 (2.0%) | 9 (2.2%) | 12 (8.9%) | 5 (6.8%) | 74 (33%) |  |
| Race not stated | 1 (<0.1%) | 0 (0%) | 0 (0%) | 0 (0%) | 0 (0%) |  |
| Unknown | 1,495 | 228 | 120 | 79 | 289 |  |
| **Stated ethnicity** |  |  |  |  |  | <0.001 |
| Not Hispanic or Latino | 1,072 (98%) | 150 (96%) | 72 (99%) | 40 (78%) | 33 (13%) |  |
| Hispanic or Latino | 22 (2.0%) | 7 (4.5%) | 1 (1.4%) | 11 (22%) | 213 (87%) |  |
| Unknown | 2,570 | 478 | 182 | 102 | 268 |  |
| **Age at specimen**<br>**collection** | 65 (58, 73) | 64 (58, 71) | 64 (58, 71) | 63 (55, 72) | 62 (57, 70) | <0.001 |
| **Age at onset** | 64 (57, 72) | 63 (57, 70) | 63 (57, 70) | 62 (55, 71) | 62 (56, 69) | <0.001 |
| **Gender** |  |  |  |  |  | 0.8 |
| Female | 1,549 (42%) | 274 (43%) | 99 (39%) | 62 (41%) | 218 (43%) |  |
| Male | 2,104 (58%) | 360 (57%) | 155 (61%) | 91 (59%) | 294 (57%) |  |
| Unknown | 11 | 1 | 1 | 0 | 2 |  |
| **xT assay version** |  |  |  |  |  | 0.8 |
| xT.v4 | 2,535 (69%) | 443 (70%) | 179 (70%) | 106 (69%) | 349 (68%) |  |
| xT.v2 | 513 (14%) | 94 (15%) | 37 (15%) | 20 (13%) | 65 (13%) |  |
| xT.v3 | 616 (17%) | 98 (15%) | 39 (15%) | 27 (18%) | 100 (19%) |  |
| **Smoking status** |  |  |  |  |  | <0.001 |
| Never smoker | 1,260 (49%) | 231 (52%) | 113 (63%) | 52 (52%) | 206 (58%) |  |
| Ever smoker | 1,325 (51%) | 212 (48%) | 66 (37%) | 48 (48%) | 150 (42%) |  |
| Unknown | 1,079 | 192 | 76 | 53 | 158 |  |
| **Cancer stage** |  |  |  |  |  | 0.6 |
| Stage 1 | 14 (0.6%) | 6 (1.4%) | 1 (0.6%) | 1 (1.0%) | 3 (0.9%) |  |
| Stage 2 | 109 (4.8%) | 20 (4.6%) | 6 (3.7%) | 5 (5.2%) | 17 (5.3%) |  |
| Stage 3 | 310 (14%) | 56 (13%) | 26 (16%) | 11 (11%) | 57 (18%) |  |
| Stage 4 | 1,850 (81%) | 352 (81%) | 129 (80%) | 79 (82%) | 246 (76%) |  |
| Unknown | 1,381 | 201 | 93 | 57 | 191 |  |
| **Tumor grade** |  |  |  |  |  | 0.01 |
| Low | 227 (9.1%) | 35 (8.2%) | 17 (8.9%) | 3 (3.2%) | 47 (13%) |  |
| Medium | 1,690 (68%) | 318 (74%) | 135 (71%) | 67 (71%) | 261 (70%) |  |
| High | 573 (23%) | 76 (18%) | 38 (20%) | 24 (26%) | 66 (18%) |  |
| Unknown | 1,174 | 206 | 65 | 59 | 140 |  |
| **MSI status** |  |  |  |  |  | 0.008 |
| Low/Stable | 3,427 (94%) | 612 (97%) | 247 (97%) | 147 (97%) | 483 (94%) |  |
| High | 233 (6.4%) | 22 (3.5%) | 8 (3.1%) | 5 (3.3%) | 31 (6.0%) |  |
| Unknown | 4 | 1 | 0 | 1 | 0 |  |
| **TMB count** | 3 (2, 5) | 4 (2, 6) | 4 (2, 5) | 3 (2, 5) | 3 (2, 5) | 0.093 |
| Unknown | 2 | 1 | 0 | 0 | 0 |  |
| **Tumor/normal tissue status** |  |  |  |  |  | 0.067 |
| Tumor and normal | 2,216 (61%) | 391 (62%) | 146 (57%) | 90 (59%) | 342 (67%) |  |
| Tumor only | 1,445 (39%) | 243 (38%) | 109 (43%) | 62 (41%) | 172 (33%) |  |
| Unknown | 3 | 1 | 0 | 1 | 0 |  |
| **Cancer primary site name** |  |  |  |  |  | 0.03 |
| Left colon | 366 (10.0%) | 59 (9.3%) | 36 (14%) | 21 (14%) | 58 (11%) |  |
| Not specified | 2,280 (62%) | 411 (65%) | 143 (56%) | 88 (58%) | 315 (61%) |  |
| Rectum | 577 (16%) | 72 (11%) | 46 (18%) | 27 (18%) | 85 (17%) |  |
| Right colon | 441 (12%) | 93 (15%) | 30 (12%) | 17 (11%) | 56 (11%) |  |
| **Tumor tissue site** |  |  |  |  |  | <0.001 |
| CRC (primary) | 1,935 (53%) | 366 (58%) | 143 (56%) | 83 (54%) | 306 (60%) |  |
| Liver | 828 (23%) | 152 (24%) | 43 (17%) | 34 (22%) | 113 (22%) |  |
| Lung | 209 (5.7%) | 21 (3.3%) | 16 (6.3%) | 2 (1.3%) | 19 (3.7%) |  |
| Other | 680 (19%) | 95 (15%) | 53 (21%) | 34 (22%) | 75 (15%) |  |
| Unknown | 12 | 1 | 0 | 0 | 1 |  |

<sup>1</sup> n (%); Median (IQR)
<sup>2</sup> Fisher's Exact Test for Count Data with simulated p-value (based on 2000 replicates) for categorical variables with any expected cell count <5; Pearson's Chi-squared test for categorical variables with all expected cell counts ≥5; Kruskal-Wallis rank sum test for continuous variables

**Supplementary Table S5.** EOCRC patient characteristics by imputed race and ethnicity group.

| Characteristic | NH White,<br>N = 1,096<sup>1</sup> | NH Black,<br>N = 241<sup>1</sup> | Asian,<br>N = 101<sup>1</sup> | Complex,<br>N = 58<sup>1</sup> | Hispanic/Latino,<br>N = 296<sup>1</sup> | p-value<sup>2</sup> |
| --- | --- | --- | --- | --- | --- | --- |
| **Stated race** |  |  |  |  |  | <0.001 |
| White | 564 (97%) | 5 (3.4%) | 2 (4.4%) | 21 (75%) | 61 (62%) |  |
| American Indian or Alaska Native | 1 (0.2%) | 0 (0%) | 0 (0%) | 2 (7.1%) | 6 (6.1%) |  |
| Asian | 0 (0%) | 0 (0%) | 39 (87%) | 1 (3.6%) | 0 (0%) |  |
| Black or African American | 3 (0.5%) | 138 (95%) | 0 (0%) | 0 (0%) | 1 (1.0%) |  |
| Native Hawaiian or Other Pacific Islander | 0 (0%) | 0 (0%) | 1 (2.2%) | 0 (0%) | 1 (1.0%) |  |
| Other Race | 10 (1.7%) | 3 (2.1%) | 3 (6.7%) | 4 (14%) | 28 (29%) |  |
| Race not stated | 1 (0.2%) | 0 (0%) | 0 (0%) | 0 (0%) | 1 (1.0%) |  |
| Unknown | 517 | 95 | 56 | 30 | 198 |  |
| **Stated ethnicity** |  |  |  |  |  | <0.001 |
| Not Hispanic or Latino | 284 (98%) | 55 (92%) | 25 (100%) | 14 (74%) | 23 (14%) |  |
| Hispanic or Latino | 7 (2.4%) | 5 (8.3%) | 0 (0%) | 5 (26%) | 137 (86%) |  |
| Unknown | 805 | 181 | 76 | 39 | 136 |  |
| **Age at specimen collection** | 45 (40, 49) | 45 (40, 49) | 45 (39, 49) | 45 (36, 48) | 44 (40, 47) | 0.056 |
| **Age at onset** | 43.9 (38.5, 47.5) | 44.3 (39.5, 47.5) | 44.1 (37.9, 47.2) | 43.3 (35.2, 46.5) | 42.8 (38.5, 46.7) | 0.12 |
| **Gender** |  |  |  |  |  | 0.033 |
| Female | 471 (43%) | 123 (51%) | 49 (49%) | 33 (57%) | 145 (49%) |  |
| Male | 623 (57%) | 118 (49%) | 51 (51%) | 25 (43%) | 151 (51%) |  |
| Unknown | 2 | 0 | 1 | 0 | 0 |  |
| **xT assay version** |  |  |  |  |  | 0.04 |
| xT.v4 | 718 (66%) | 166 (69%) | 70 (69%) | 44 (76%) | 222 (75%) |  |
| xT.v2 | 167 (15%) | 28 (12%) | 17 (17%) | 7 (12%) | 26 (8.8%) |  |
| xT.v3 | 211 (19%) | 47 (20%) | 14 (14%) | 7 (12%) | 48 (16%) |  |
| **Smoking status** |  |  |  |  |  | 0.077 |
| never_smoker | 464 (62%) | 90 (61%) | 53 (79%) | 23 (66%) | 132 (65%) |  |
| ever_smoker | 282 (38%) | 58 (39%) | 14 (21%) | 12 (34%) | 70 (35%) |  |
| Unknown | 350 | 93 | 34 | 23 | 94 |  |
| **Cancer stage** |  |  |  |  |  | 0.025 |
| Stage 1 | 5 (0.8%) | 0 (0%) | 0 (0%) | 1 (2.7%) | 2 (1.1%) |  |
| Stage 2 | 17 (2.6%) | 4 (2.8%) | 1 (1.8%) | 4 (11%) | 5 (2.8%) |  |
| Stage 3 | 73 (11%) | 23 (16%) | 10 (18%) | 7 (19%) | 36 (20%) |  |
| Stage 4 | 549 (85%) | 116 (81%) | 44 (80%) | 25 (68%) | 133 (76%) |  |
| Unknown | 452 | 98 | 46 | 21 | 120 |  |
| **Tumor grade** |  |  |  |  |  | 0.14 |
| Low | 71 (9.8%) | 10 (6.0%) | 10 (14%) | 4 (11%) | 20 (9.8%) |  |
| Medium | 519 (72%) | 118 (70%) | 44 (63%) | 21 (58%) | 153 (75%) |  |
| High | 135 (19%) | 40 (24%) | 16 (23%) | 11 (31%) | 32 (16%) |  |
| Unknown | 371 | 73 | 31 | 22 | 91 |  |
| **MSI status** |  |  |  |  |  | 0.074 |
| Low/Stable | 1,057 (97%) | 228 (95%) | 98 (97%) | 54 (93%) | 278 (94%) |  |
| High | 35 (3.2%) | 13 (5.4%) | 3 (3.0%) | 4 (6.9%) | 18 (6.1%) |  |
| Unknown | 4 | 0 | 0 | 0 | 0 |  |
| **TMB count** | 3.1 (1.9, 4.2) | 3.8 (2.6, 5.9) | 3.3 (2.3, 5.1) | 3.1 (2.3, 5.0) | 3.1 (1.9, 4.6) | <0.001 |
| Unknown | 0 | 1 | 1 | 0 | 0 |  |
| **Tumor/normal tissue status** |  |  |  |  |  | 0.7 |
| Tumor and normal | 691 (63%) | 152 (63%) | 60 (59%) | 38 (66%) | 198 (67%) |  |
| Tumor only | 402 (37%) | 89 (37%) | 41 (41%) | 20 (34%) | 98 (33%) |  |
| Unknown | 3 | 0 | 0 | 0 | 0 |  |
| **Cancer primary site name** |  |  |  |  |  | 0.087 |
| Left colon | 113 (10%) | 22 (9.1%) | 13 (13%) | 5 (8.6%) | 32 (11%) |  |
| Not specified | 719 (66%) | 148 (61%) | 63 (62%) | 32 (55%) | 179 (60%) |  |
| Rectum | 197 (18%) | 46 (19%) | 17 (17%) | 15 (26%) | 72 (24%) |  |
| Right colon | 67 (6.1%) | 25 (10%) | 8 (7.9%) | 6 (10%) | 13 (4.4%) |  |
| **Tumor tissue site** |  |  |  |  |  | 0.016 |
| CRC (primary) | 561 (51%) | 124 (51%) | 43 (43%) | 29 (51%) | 174 (59%) |  |
| Liver | 249 (23%) | 48 (20%) | 20 (20%) | 10 (18%) | 48 (16%) |  |
| Lung | 74 (6.8%) | 11 (4.6%) | 14 (14%) | 7 (12%) | 19 (6.4%) |  |
| Other | 206 (19%) | 58 (24%) | 24 (24%) | 11 (19%) | 54 (18%) |  |
| Unknown | 6 | 0 | 0 | 1 | 1 |  |

<sup>1</sup> n (%); Median (IQR)
<sup>2</sup> Fisher's Exact Test for Count Data with simulated p-value (based on 2000 replicates) for categorical variables with any expected cell count <5; Pearson's Chi-squared test for categorical variables with all expected cell counts ≥5; Kruskal-Wallis rank sum test for continuous variables

**Supplementary Table S6:**
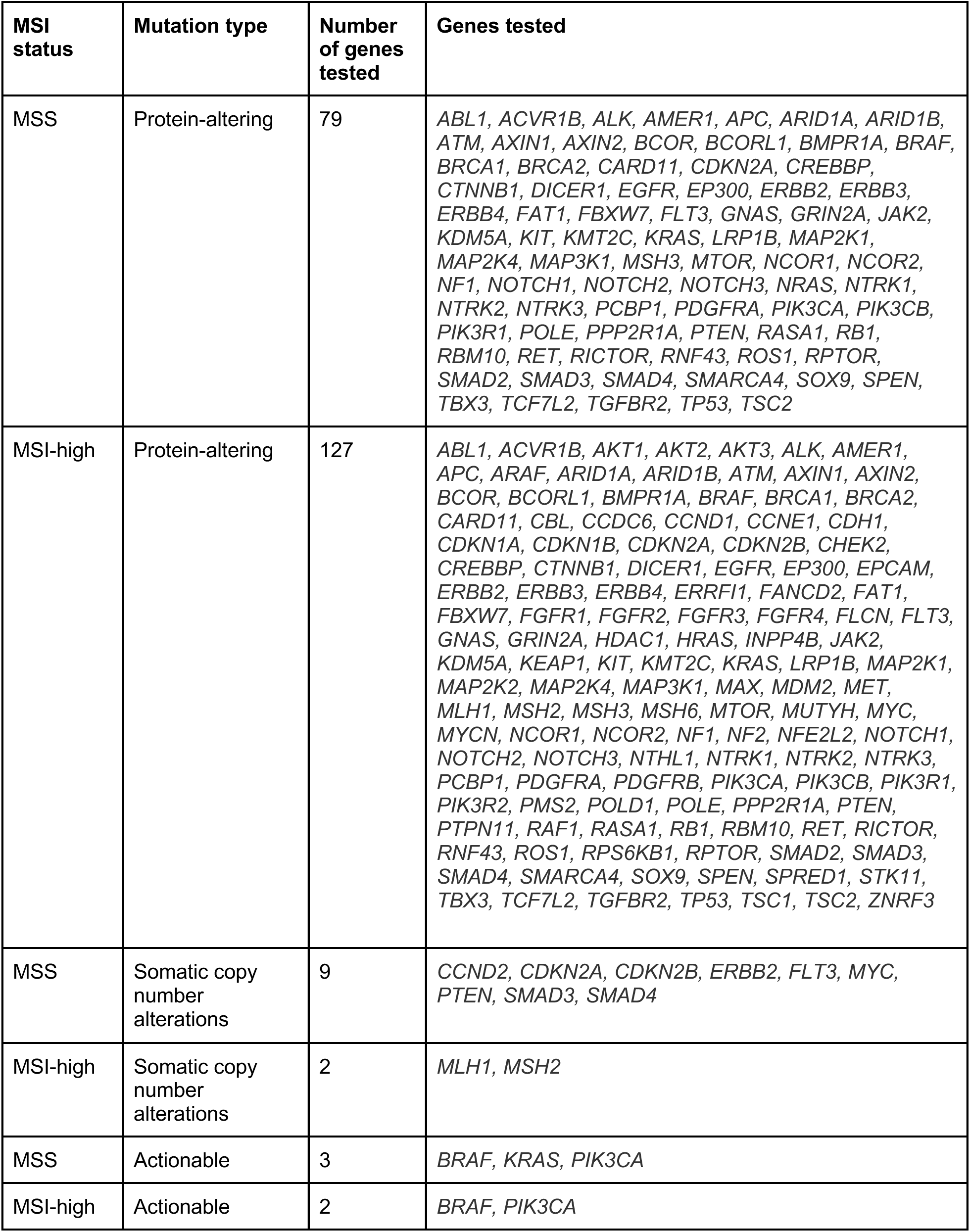
Genes tested by mutation type and MSI status.

**Supplementary Table S7:** Somatic mutation associations with imputed race and ethnicity in MSS patients that differ by onset age or primary site. Mutation type = type of mutation tested. “SCNA” refers to somatic copy number alterations, and “actionable” refers to protein-altering mutations that are classified with a OncoKB Therapeutic Level of Evidence V2 designation therapeutic level 1 or 2, or resistance level R1, irrespective of the solid cancer type. Gene = gene symbol of tested gene. N total = total number of patients included in models. N with mutation = number of patients included in models who have one or more of the mutation type in the gene. *P* LR (FDR) = *P*-value for the likelihood ratio test, adjusted to control the false discovery rate. Intxn. imputed r/e group = imputed race and ethnicity group with differing association by either onset age or primary site with the presence of mutations in this gene in logistic regression test (i.e., the group that shows an interaction effect). Intxn. group = the specific onset group or primary site for which an OR is estimated. OR (95% CI) = odds ratio compared to NH White category and 95% confidence interval for the given ancestry proportion in the logistic regression test. *P* Logistic = *P*-value for the specific ancestry proportion in the logistic regression test, not adjusted for multiple tests.

| Mutation type | Gene | N total | N with mutation | *P* LR (FDR) | Intxn. imputed r/e group | *P* int. term | Intxn. group | OR (95% CI) | *P* Logistic |
| --- | --- | --- | --- | --- | --- | --- | --- | --- | --- |
| SCNA | *FLT3* | 6,413 | 199 | 0.033 | Hispanic /Latino | 0.002 | AOCRC | 2.38 (1.53, 3.72) | 1.4e-04 |
|  |  |  |  |  |  |  | EOCRC | 0.46 (0.18, 1.18) | 0.107 |
| Actionable | *BRAF* | 2,604 | 116 | 0.005 | NH Asian | 0.031 | Colon | 0.12 (0.16, 0.86) | 0.035 |
|  |  |  |  |  |  |  | Rectum | 1.84 (0.41, 8.32) | 0.426 |
|  |  |  |  |  | NH Black | 0.001 | Colon | 0.10 (0.03, 0.42) | 0.002 |
|  |  |  |  |  |  |  | Rectum | 2.12 (0.68, 6.59) | 0.192 |

**Supplementary Table S8:** Somatic mutation associations with imputed race and ethnicity in MSI-high patients. Mutation type = type of mutation tested. “Non-silent” refers to protein-altering mutations and “SCNA” refers to somatic copy number alterations. Gene = gene symbol of tested gene. N total = total number of patients included in models. N with mutation = number of patients included in models who have one or more of the mutation type in the gene. *P* LR (FDR) = *P*-value for the likelihood ratio test, adjusted to control the false discovery rate. Imputed race and ethnicity group = imputed race and ethnicity group associated with the presence of mutations in this gene in logistic regression test. OR (95% CI) = odds ratio compared to NH White category and 95% confidence interval for the given ancestry proportion in the logistic regression test. *P* Logistic = *P*-value for the specific ancestry proportion in the logistic regression test, not adjusted for multiple tests.

| Mutation type | Gene | N total | N with mutation | *P* LR (FDR) | Imputed race and ethnicity group | OR (95% CI) | *P* Logistic |
| --- | --- | --- | --- | --- | --- | --- | --- |
| Protein-altering | *KMT2C* | 270 | 153 | 0.005 | NH Black | 23.7 (3.1, 181) | 0.002 |
| SCNA | *MLH1* | 432 | 13 | 0.005 | NH Asian | 13.9 (1.9, 103) | 0.01 |
|  |  |  |  |  | Hispanic/Latino | 11.4 (2.6, 49.6) | 0.001 |

**Supplementary Table S9:** Patients characteristics for those patients included in RNA-Seq analyses.

|  | **Colon/Rectum**<br>**(N=1957)** | **Liver**<br>**(N=788)** |
| --- | --- | --- |
| **Imputed race and ethnicity category** |  |  |
| NH Black | 252 (12.9%) | 112 (14.2%) |
| NH Asian | 98 (5.0%) | 29 (3.7%) |
| Complex | 66 (3.4%) | 20 (2.5%) |
| NH White | 1254 (64.1%) | 544 (69.0%) |
| Hispanic/Latino | 287 (14.7%) | 83 (10.5%) |
| **Age at diagnosis** |  |  |
| Mean (SD) | 57.8 (13.6) | 57.3 (12.8) |
| Median [Min, Max] | 57.8 [1.39, 88.7] | 56.6 [16.2, 89.1] |
| **Early or Average Onset** |  |  |
| Early | 643 (32.9%) | 239 (30.3%) |
| Average | 1314 (67.1%) | 549 (69.7%) |
| **Gender** |  |  |
| Female | 846 (43.2%) | 318 (40.4%) |
| Male | 1111 (56.8%) | 470 (59.6%) |
| **RNA assay version** |  |  |
| RS | 658 (33.6%) | 259 (32.9%) |
| RS.v2 | 1299 (66.4%) | 529 (67.1%) |
| **Stage** |  |  |
| 1 | 16 (0.8%) | 0 (0%) |
| 2 | 88 (4.5%) | 0 (0%) |
| 3 | 289 (14.8%) | 1 (0.1%) |
| 4 | 786 (40.2%) | 544 (69.0%) |
| unknown | 778 (39.8%) | 243 (30.8%) |
| **Grade** |  |  |
| Grade 1 | 133 (6.8%) | 33 (4.2%) |
| Grade 2 | 1083 (55.3%) | 320 (40.6%) |
| Grade 3 | 333 (17.0%) | 83 (10.5%) |
| Unknown | 408 (20.8%) | 352 (44.7%) |
| **MSI Status** |  |  |
| High | 127 (6.5%) | 10 (1.3%) |
| Stable | 1830 (93.5%) | 778 (98.7%) |
| **Tumor tissue Site** |  |  |
| Colon | 1279 (65.4%) | 0 (0%) |
| Rectum | 678 (34.6%) | 0 (0%) |
| Liver | 0 (0%) | 788 (100%) |

**Supplementary Table S10:** Genes significantly under or over expressed in gene sets found significant. The “TMM” method corresponds to the inputs to mROAST and follows the workflow: Prevalence filtering -> TMM -> VOOM -> LIMMA linear model with eBayes and batch as a covariate. The “VST” method corresponds to the inputs to GSVA and follows the workflow: Prevalence filtering -> VST -> removeBatchEffects -> LIMMA linear model with eBayes. Genes reported here were required to have a p value <0.05 in at least one of the above workflows. All results reported in this table for the Non-Hispanic Black imputed category are in comparison to Non-Hispanic White; all results for AFR represent changes in gene expression as the dominance of AFR compared to other ancestries increases. Genes are presented ranked by decreasing fold change by the TMM method.

| **S10.1: NH Black Colon/Rectum** |  |  |  |  |  |
| --- | --- | --- | --- | --- | --- |
| **Gene** | **Log2 Fold change TMM** | **P Value TMM** | **Log2 fold change VST** | **P Value VST** | **Gene Set** |
| *SERPINC1* | 0.674 | <0.001 | 0.167 | <0.001 | Hallmark Coagulation |
| *LEFTY2* | 0.501 | <0.001 | 0.273 | <0.001 | Hallmark Coagulation |
| *HMGCS2* | 0.293 | 0.057 | 0.281 | 0.027 | Hallmark Coagulation |
| *HRG* | 0.254 | 0.029 | 0.031 | 0.29 | Hallmark Coagulation |
| *ACOX2* | 0.193 | 0.002 | 0.173 | 0.001 | Hallmark Coagulation |
| *MASP2* | 0.116 | 0.053 | 0.089 | 0.042 | Hallmark Coagulation |
| *CD9* | 0.097 | 0.027 | 0.089 | 0.029 | Hallmark Coagulation |
| *LGMN* | 0.096 | 0.013 | 0.083 | 0.024 | Hallmark Coagulation |
| *LTA4H* | 0.09 | 0.012 | 0.082 | 0.014 | Hallmark Coagulation |
| *CASP9* | 0.075 | 0.012 | 0.06 | 0.018 | Hallmark Coagulation |
| *USP11* | -0.074 | 0.015 | -0.078 | 0.009 | Hallmark Coagulation |
| *FYN* | -0.089 | 0.049 | -0.089 | 0.043 | Hallmark Coagulation |
| *BMP1* | -0.091 | 0.03 | -0.093 | 0.025 | Hallmark Coagulation |
| *CAPN2* | -0.107 | 0.003 | -0.11 | 0.001 | Hallmark Coagulation |
| *SLC25A18* | -0.109 | 0.066 | -0.049 | 0.034 | BioCarta Rhodopsin |
| *CTSH* | -0.109 | 0.008 | -0.112 | 0.005 | Hallmark Coagulation |
| *CFP* | -0.122 | 0.03 | -0.059 | 0.09 | BioCarta Alternative |
| *CRIP2* | -0.122 | 0.017 | -0.087 | 0.026 | Hallmark Coagulation |
| *LRP1* | -0.132 | 0.008 | -0.135 | 0.005 | Hallmark Coagulation |
| *RCVRN* | -0.134 | 0.076 | -0.039 | 0.042 | BioCarta Rhodopsin |
| *SPARC* | -0.138 | 0.036 | -0.143 | 0.029 | Hallmark Coagulation |
| *CSRP1* | -0.14 | 0.011 | -0.141 | 0.009 | Hallmark Coagulation |
| *THBS1* | -0.141 | 0.024 | -0.146 | 0.02 | Hallmark Coagulation |
| *TIMP1* | -0.144 | 0.017 | -0.118 | 0.039 | Hallmark Coagulation,<br>BioCarta RECK |
| *PLAU* | -0.147 | 0.033 | -0.118 | 0.057 | Hallmark Coagulation |
| *PDE6B* | -0.156 | 0.004 | -0.084 | 0.002 | BioCarta Rhodopsin |
| *FBN1* | -0.165 | 0.02 | -0.163 | 0.02 | Hallmark Coagulation |
| *PDE6A* | -0.176 | 0.048 | -0.064 | 0.049 | BioCarta Rhodopsin |
| *ITGB3* | -0.177 | <0.001 | -0.162 | <0.001 | Hallmark Coagulation |
| *MMP14* | -0.178 | 0.001 | -0.18 | 0.001 | Hallmark Coagulation,<br>BioCarta RECK |
| *VWF* | -0.181 | 0.001 | -0.184 | 0.001 | Hallmark Coagulation |
| *MMP2* | -0.184 | 0.012 | -0.19 | 0.008 | Hallmark Coagulation,<br>BioCarta RECK |
| *F8* | -0.186 | 0.001 | -0.127 | 0.001 | Hallmark Coagulation |
| *F2* | -0.194 | 0.033 | -0.074 | 0.03 | Hallmark Coagulation |
| *TIMP2* | -0.203 | 0.001 | -0.204 | <0.001 | BioCarta RECK |
| *CFI* | -0.204 | 0.002 | -0.201 | 0.001 | Hallmark Coagulation |
| *SERPING1* | -0.208 | 0.002 | -0.211 | 0.001 | Hallmark Coagulation |
| *CFH* | -0.209 | 0.008 | -0.207 | 0.005 | Hallmark Coagulation |
| *RGN* | -0.215 | 0.018 | -0.083 | 0.018 | Hallmark Coagulation |
| *F2RL2* | -0.218 | 0.002 | -0.169 | 0.001 | Hallmark Coagulation |
| *C1R* | -0.22 | <0.001 | -0.223 | <0.001 | Hallmark Coagulation |
| *TIMP4* | -0.224 | 0.001 | -0.098 | 0.003 | BioCarta RECK |
| *DPP4* | -0.232 | 0.012 | -0.204 | 0.011 | Hallmark Coagulation |
| *GSN* | -0.233 | <0.001 | -0.235 | <0.001 | Hallmark Coagulation |
| *TMPRSS6* | -0.235 | <0.001 | -0.104 | 0.001 | Hallmark Coagulation |
| *C1S* | -0.238 | <0.001 | -0.239 | <0.001 | Hallmark Coagulation |
| *FN1* | -0.246 | 0.006 | -0.251 | 0.005 | Hallmark Coagulation |
| *HTRA1* | -0.247 | <0.001 | -0.217 | 0.001 | Hallmark Coagulation |
| *S100A13* | -0.248 | <0.001 | -0.221 | <0.001 | Hallmark Coagulation |
| *S100A1* | -0.251 | <0.001 | -0.135 | <0.001 | Hallmark Coagulation |
| *TF* | -0.256 | 0.003 | -0.12 | 0.011 | Hallmark Coagulation |
| *CTSK* | -0.279 | <0.001 | -0.272 | <0.001 | Hallmark Coagulation |
| *TIMP3* | -0.308 | <0.001 | -0.275 | <0.001 | Hallmark Coagulation,<br>BioCarta RECK |
| *MMP11* | -0.31 | 0.001 | -0.302 | 0.001 | Hallmark Coagulation |
| *HPN* | -0.319 | 0.02 | -0.166 | 0.007 | Hallmark Coagulation |
| *MMP7* | -0.367 | 0.012 | -0.302 | 0.008 | Hallmark Coagulation |
| *C3* | -0.378 | <0.001 | -0.379 | <0.001 | Hallmark Coagulation,<br>BioCarta Alternative |
| *F11* | -0.38 | 0.002 | -0.104 | 0.002 | Hallmark Coagulation |
| *MMP8* | -0.382 | 0.008 | -0.106 | 0.016 | Hallmark Coagulation |
| *MMP3* | -0.428 | 0.006 | -0.37 | 0.004 | Hallmark Coagulation |
| *CFB* | -0.52 | <0.001 | -0.403 | <0.001 | Hallmark Coagulation,<br>BioCarta Alternative |
| *MMP1* | -0.521 | 0.002 | -0.474 | 0.001 | Hallmark Coagulation |
| *COMP* | -0.526 | 0.008 | -0.342 | 0.024 | Hallmark Coagulation |
| *CPN1* | -0.605 | 0.001 | -0.157 | 0.001 | Hallmark Coagulation |
| *CTSE* | -0.615 | 0.002 | -0.383 | <0.001 | Hallmark Coagulation |
| **S10.2 AFR Liver** |  |  |  |  |  |
| **Gene** | **Log2 Fold**<br>**change**<br>**TMM** | **P Value**<br>**TMM** | **Log2 fold**<br>**change VST** | **P Value**<br>**VST** | **Gene Set** |
| *FHL5* | -0.063 | <0.001 | -0.021 | <0.001 | BioCarta CREM |
| *FSHR* | -0.056 | 0.003 | -0.012 | 0.001 | BioCarta CREM |
| *ADCY1* | -0.027 | 0.048 | -0.019 | 0.088 | BioCarta CREM |

**Supplementary Table S12:**
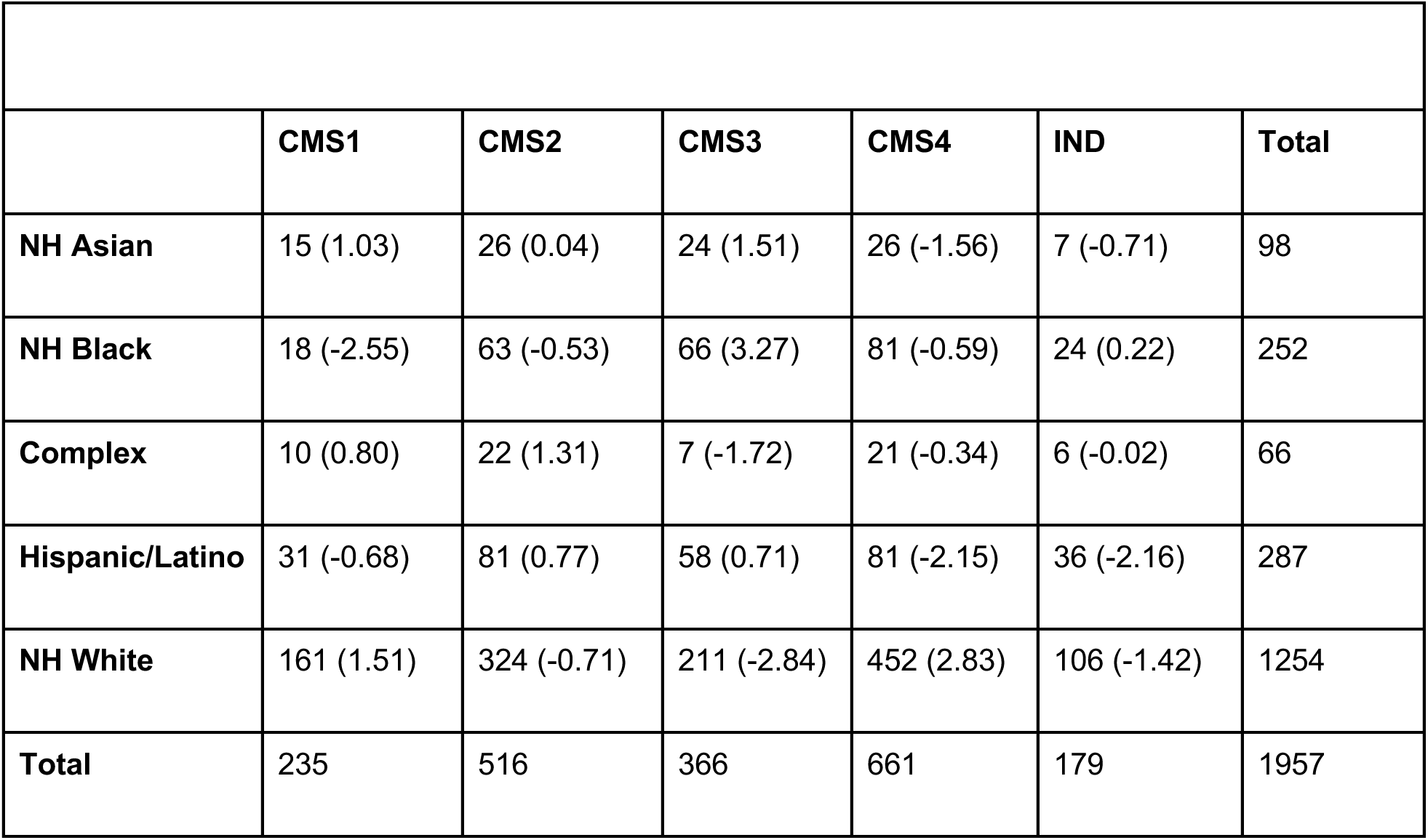
Contingency table of CMS classification and imputed racial and ethnic category. Included in parenthesis are standardized residuals from a chi-square test of independence. Absolute standardized residuals >1.96 indicate significance at α = 0.05, with the sign of the residual indicating greater than expected count (+) or less than expected count (-).

**Supplementary Table S13:**
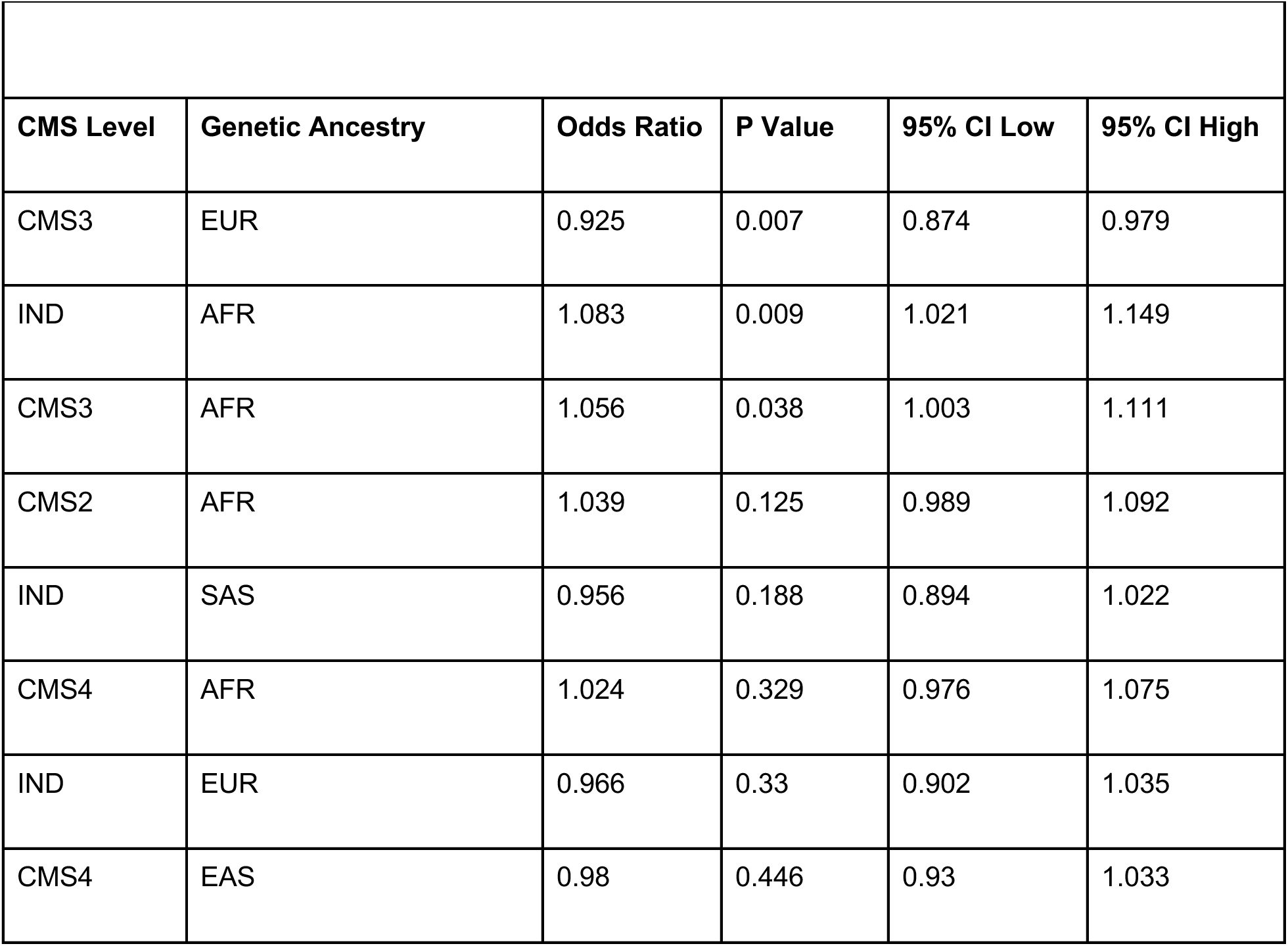
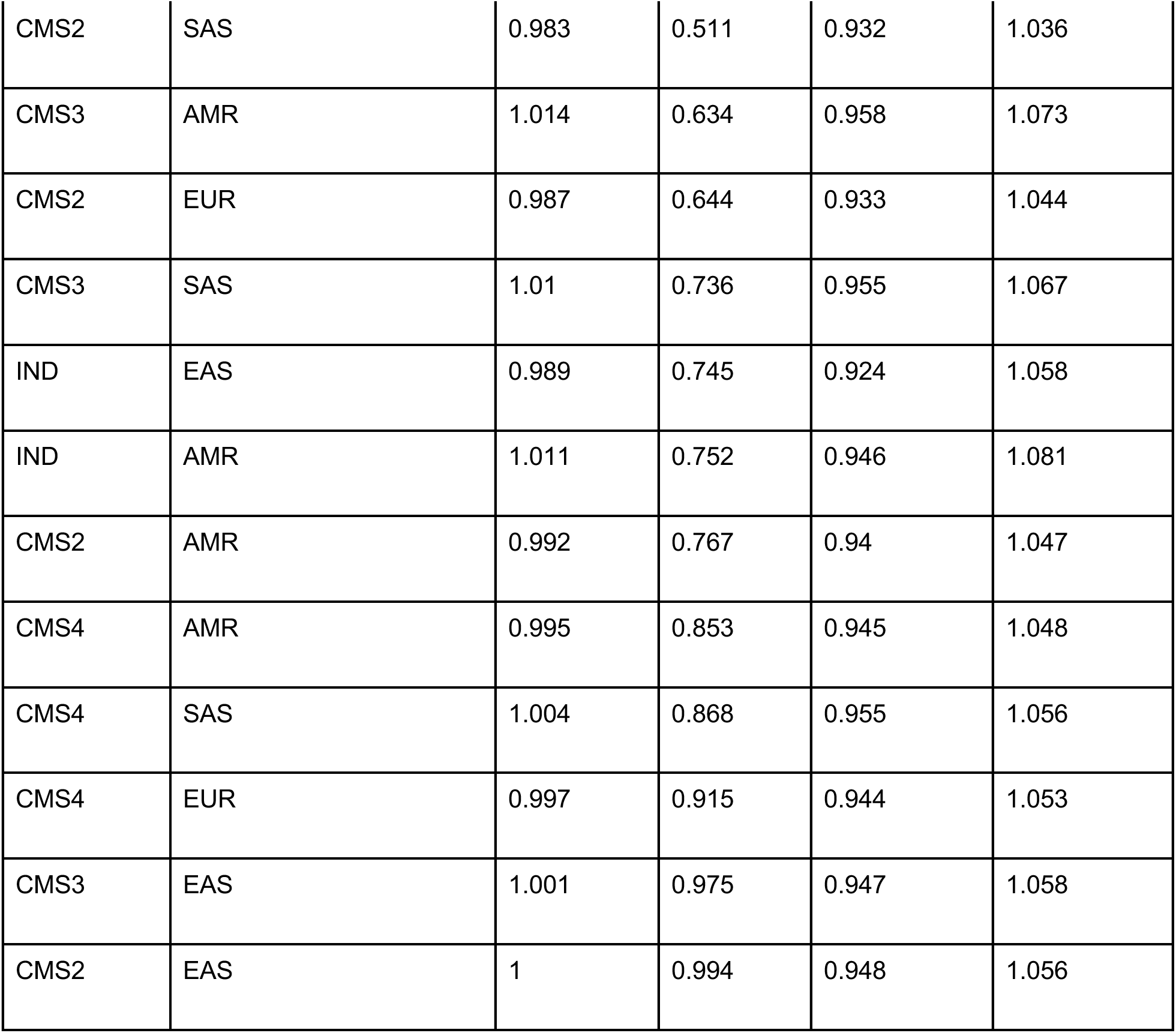
Multinomial logistic regression for CMS and genetic ancestry proportions. Odds ratios ranked by ascending p value, CMS1 as reference category.

**Supplementary Figure S1:**
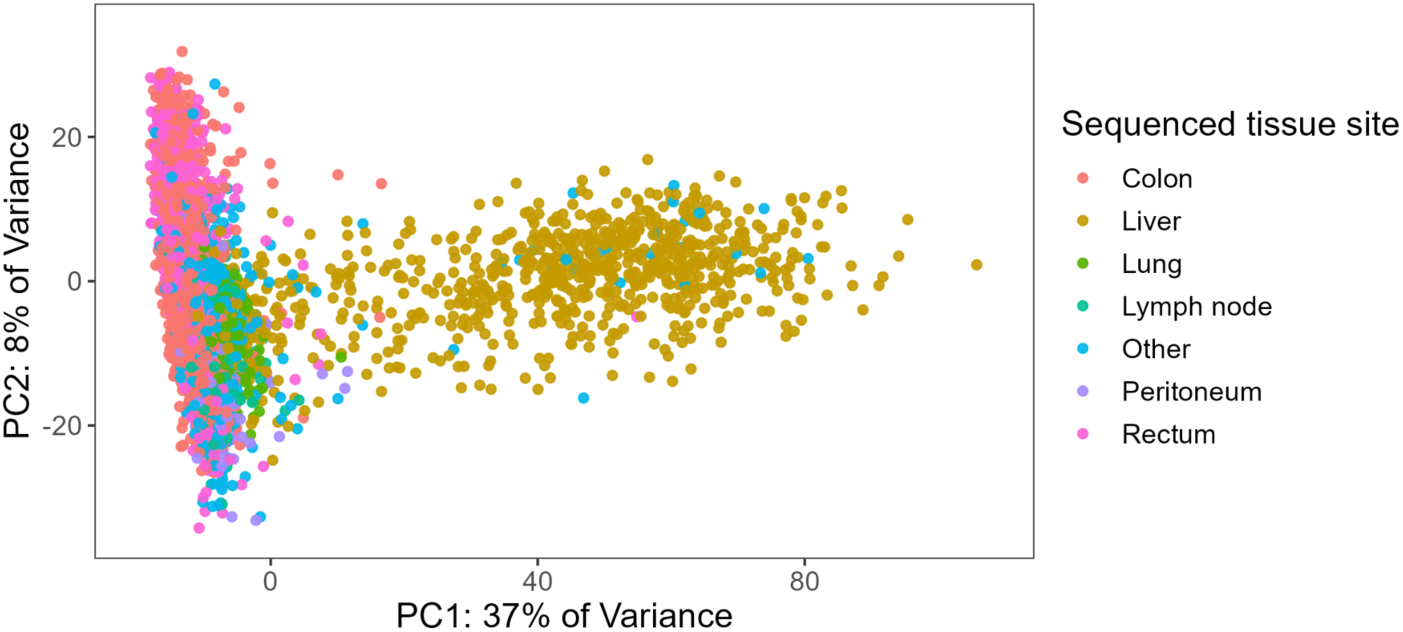
PCA Plots of expression data by sequenced tissue site

**Supplementary Figure S2:**
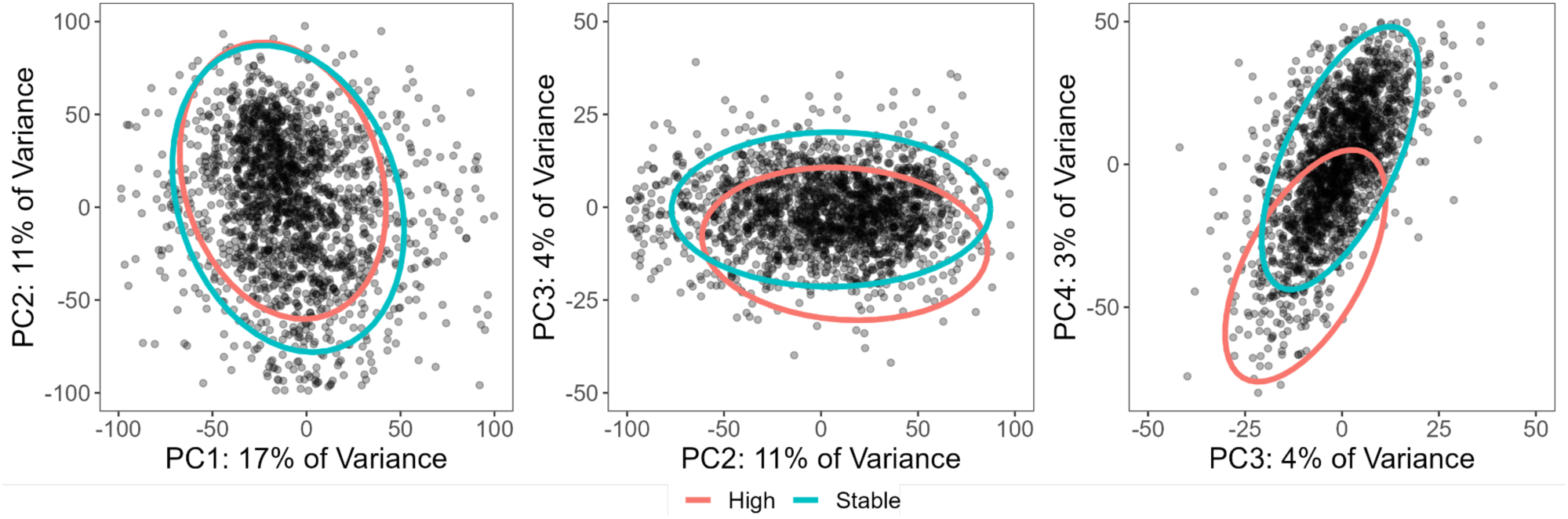
PCA Plots of expression data from only colon and rectum sequencing sites with 95% confidence interval ellipses colored by MSI Status (Multivariate normal distribution). Darker areas indicate a higher density of points.

**Supplementary figure S3:**
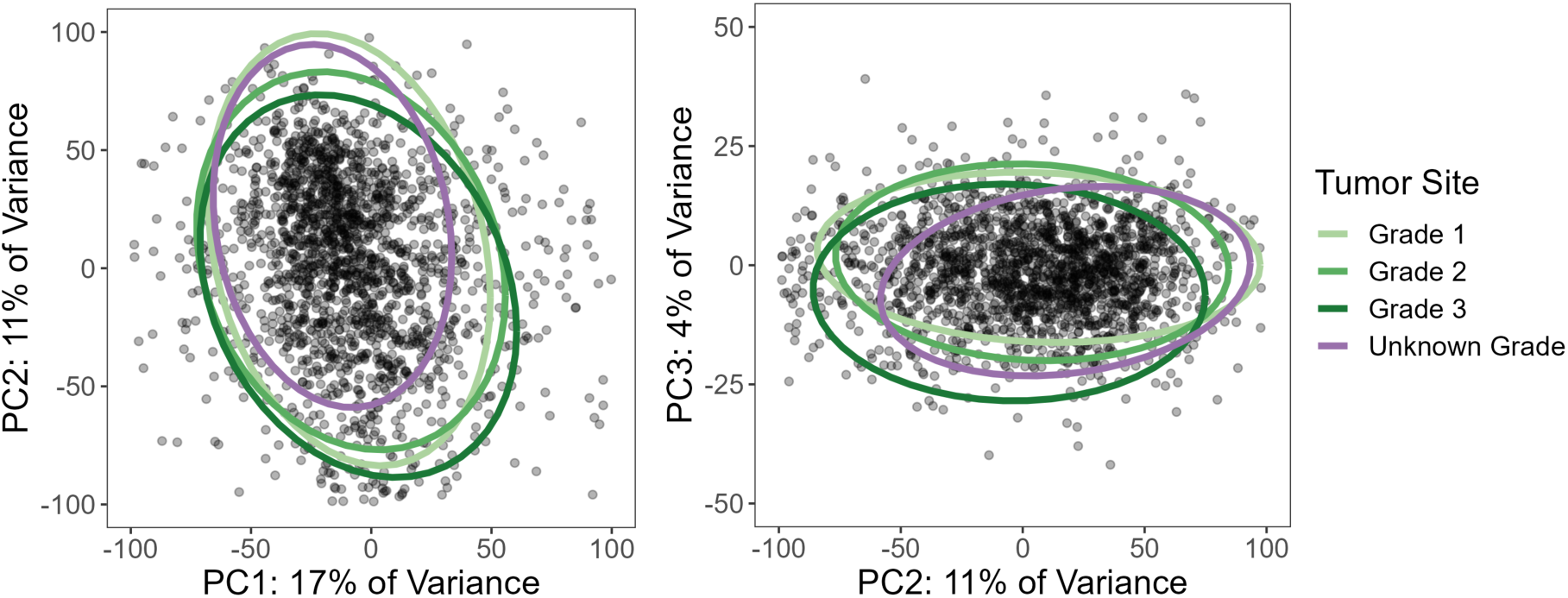
PCA Plot of expression data from colon and rectum sequencing site only with 95% confidence interval ellipses colored by tumor grade (Multivariate normal distribution)

**Supplementary Figure S4:**
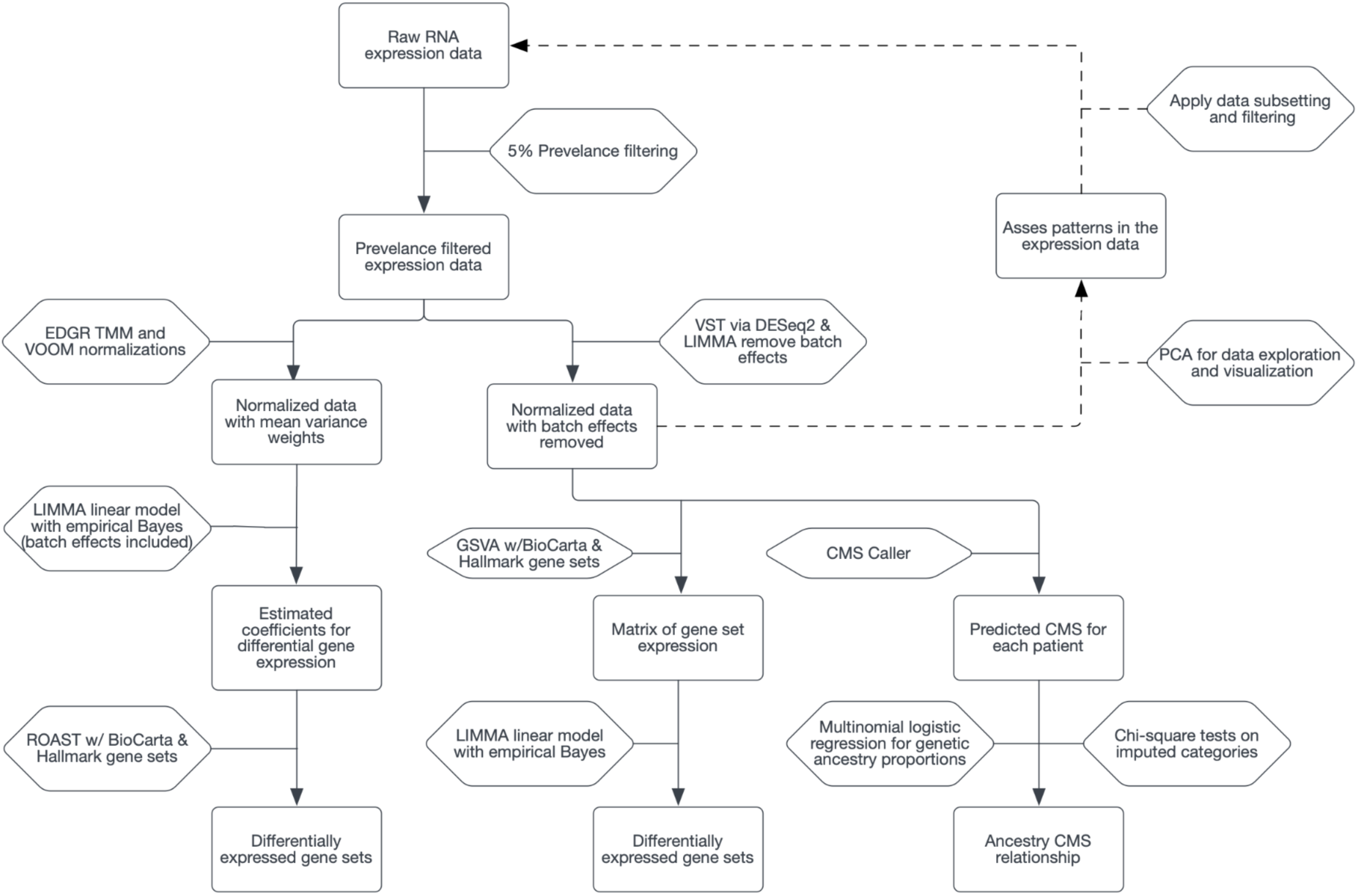
RNA analysis workflow diagram.

